# Hormone Replacement Therapy and Disability Progression in Women with Multiple Sclerosis

**DOI:** 10.64898/2026.10.21.26366096

**Authors:** Timothy P. Copeland, Kyla A. McKay, Astrid Pedersen, Roxane Handal-Orefice, Katharina Fink

## Abstract

**Background:** Evidence on menopausal hormone replacement therapy (HRT) and disability progression in women with multiple sclerosis (MS) is limited, particularly in the era of high-efficacy disease-modifying therapy. We estimated the association between systemic HRT and sustained disability progression and recurring relapses in a contemporary national cohort.

**Methods:** In a nationwide Swedish registry-based cohort, we followed women with MS aged 40 to 60 years entering from 2006 through 2024, with follow-up through 2025, under a new-user design. Systemic HRT was modeled as time-varying current use and quarterly oral-equivalent dose categories. Outcomes were sustained Expanded Disability Status Scale (EDSS) 4.0, sustained EDSS 6.0, and recurring relapses. Cox models adjusted for baseline covariates and time-varying age and disease-modifying therapy (DMT) class.

**Results:** Among 7,623 women, 1,192 (15.6%) initiated HRT. Current HRT use was associated with a lower hazard of sustained EDSS 4.0 (adjusted hazard ratio [aHR] 0.70, 95% confidence interval [CI] 0.51 to 0.96) but not sustained EDSS 6.0 (aHR 0.80, 95% CI 0.55 to 1.16) or recurring relapses (aHR 1.00, 95% CI 0.74 to 1.36). Dose categories showed no consistent gradient. Raising the cohort-entry age floor to 50 years gave a consistent EDSS 4.0 estimate (aHR 0.62, 95% CI 0.40 to 0.95).

**Conclusions:** HRT was not associated with increased relapse risk or severe disability progression. The lower hazard of sustained EDSS 4.0 was not mirrored at EDSS 6.0 and showed no dose gradient. Given menopausal status was unmeasured, replication in cohorts with verified menopausal status is required for clear clinical interpretation.

## Introduction

Multiple sclerosis (MS) disproportionately affects women, with female-to-male ratios exceeding 2:1 in most populations^1^. As a chronic disease typically diagnosed in early adulthood^2^, most women with MS will reach menopause while living with the disease. Endogenous sex hormones modulate MS inflammatory activity, as is demonstrated by relapse rates declining during late pregnancy and rebounding postpartum^3^. Preclinical work also demonstrates that estrogen and progesterone exert immunomodulatory and neuroprotective effects through distinct pathways^4–6^. These observations have sustained interest in whether exogenous hormones may alter MS disease course^7^.

During menopause, ovarian function declines, leading to a marked drop in estrogen and near-complete cessation of progesterone production. Observational studies suggest that menopause may represent an inflection point in disability accumulation. Several cohorts have exhibited accelerated worsening after menopause, even as relapse rates declined^8–10^, a pattern confirmed by meta-analysis^11^ and by a recent study linking menopause to worse objective functional outcomes and higher serum neurofilament light chain levels^12^. Disentangling menopausal effects from age-related progression in the disease-modifying therapy (DMT) era remains difficult, and the literature shows substantial heterogeneity across studies^13,14^. Because menopausal status is not recorded in Swedish registers, a comparison of hormone replacement therapy (HRT) users to non-users contrasts women with and without dispensed HRT rather than estrogen-replete versus estrogen-depleted women, and is best interpreted in terms of HRT use rather than menopausal estrogen status.

However, direct evidence on exogenous HRT in MS is limited^15^. The only prior population-based cohort study, from Denmark, found no evidence that HRT protects against disability accrual and reported a 20% higher rate of recurrent relapses among current users (hazard ratio 1.2, 95% CI 1.0 to 1.4), which the authors attributed to the earliest calendar period (1996 to 2005)^16^. By design or study period, most prior studies do not reflect the contemporary distribution of DMTs, which are associated with better long-term disability outcomes^17^. A recent review concluded that available data are insufficient to support or discourage HRT in MS^18^, and a published debate illustrated the clinical uncertainty: Voskuhl^19^ argued that all women with MS should start HRT at menopause unless contraindicated, while Magyari^20^ countered that the evidence base does not yet support universal recommendation.

We aimed to estimate the association between systemic HRT exposure and disability progression in a contemporary nationwide cohort where high-efficacy DMTs are widely used. Primary outcomes were sustained progression to Expanded Disability Status Scale (EDSS) 4.0 and 6.0, representing meaningful mobility impairment and need for walking assistance, respectively^21^, and recurring relapses.

## Methods

### Study Design

We conducted a nationwide registry-based cohort study of women with multiple sclerosis (MS) in Sweden, using a new-user design (i.e., no systemic HRT at cohort entry) and modeling HRT as a time-varying exposure with both a binary exposed/unexposed indicator and quarterly oral-equivalent dose categories. The study follows the RECORD extension of STROBE^22,23^.

### Data Sources

Data were obtained by linking the Swedish MS Registry (SMSreg)^24^ to national registries via the personal identification number assigned to all Swedish residents. SMSreg captures EDSS assessments (i.e., score and date), MS phenotype, relapse dates, and DMT episodes. The Prescribed Drug Register^25^ provided HRT data from dispensed prescriptions available since July 2005, and the National Patient Register^26^ provided ICD-10 diagnoses and procedure codes for exclusions and covariates. Additional sources were the Swedish Cancer Registry^27^, the Longitudinal Integrated Database for Health Insurance and Labour Market Studies (LISA)^28^, the Register of the Total Population^29^, the Medical Birth Registry^30^, the Migration Register^31^, and the Cause of Death Register^32^, all with established validity (Appendix A).

### Study Population

The study period was January 1, 2006, to December 31, 2025. We included women with an MS diagnosis recorded in SMSreg who were aged 40 to 60 years at cohort entry. The age window targets the perimenopausal and early postmenopausal period when HRT decisions are most clinically relevant.

Further inclusion criteria were being alive and resident in Sweden at cohort entry, at least two EDSS assessments, at least one DMT record (i.e., a specific DMT or a documented no-treatment record), a recorded MS phenotype at diagnosis (i.e., relapsing-remitting or primary progressive; secondary progressive is not a diagnosis-onset phenotype in SMSreg), and a recorded diagnosis date.

We excluded women with any malignant cancer other than non-melanoma skin cancer before cohort entry, and women with bilateral oophorectomy, venous thromboembolism, or stroke on or before cohort entry. Women who had undergone hematopoietic stem cell transplantation or received mitoxantrone before cohort entry were also excluded. We further excluded women with any systemic HRT dispensing before cohort entry (i.e., new-user design). Women with a recorded birth after cohort entry were censored at estimated conception (i.e., 270 days before the birth date); those whose estimated conception fell on or before cohort entry were excluded entirely. For each disability outcome the at-risk population required baseline EDSS below the threshold (i.e., below 4.0 and below 6.0, respectively); the recurring-relapse analysis used the full cohort. Exclusion rationales appear in Appendix B.4 and the sequential process in Figure 1.

**Figure 1.**
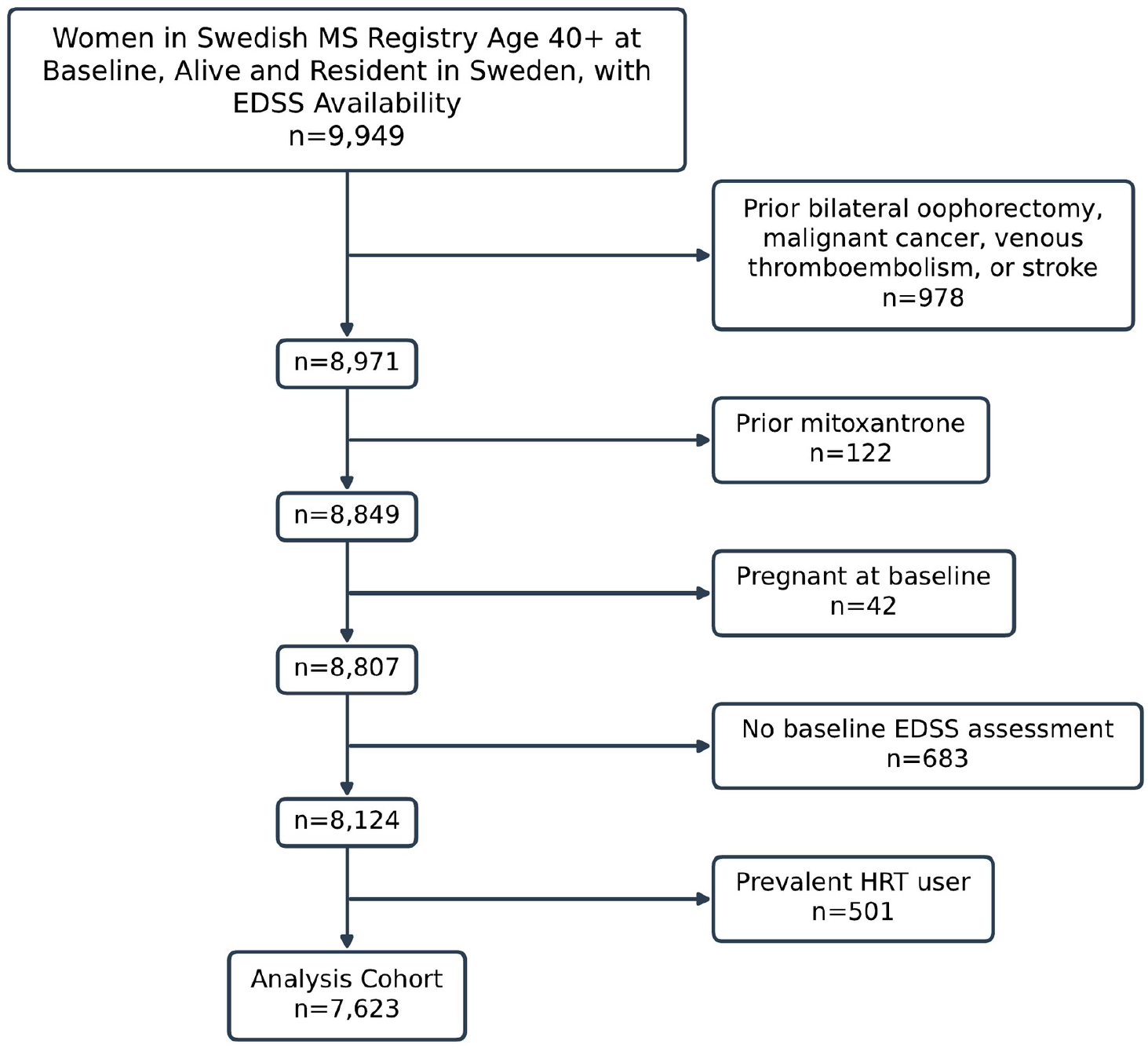
Sequential exclusion flowchart, starting from women alive and resident in Sweden at cohort entry (primary age 40+ cohort).

Cohort entry was defined as the latest of age 40, MS diagnosis date, or January 1, 2006, with entry limited to December 31, 2024, to ensure at least one year of potential follow-up. When diagnosis date was missing, the earliest recorded treatment start date was used as a proxy (Appendix B.1). Follow-up was censored at the earliest of bilateral oophorectomy, venous thromboembolism, stroke, death, emigration, or December 31, 2025.

### Exposure Definition

Systemic HRT exposure was ascertained from the Prescribed Drug Register using ATC codes for estradiol, estriol, combined estrogen-progestogen products, and tibolone, excluding vaginal formulations. Dispensings across routes were converted to a common oral estradiol-equivalent daily dose using published route-specific potency factors^33^, and the average daily dose was aggregated within 91-day exposure quarters. Because Swedish guidelines require a progestogen add-on for women with an intact uterus, given the cancer risk of unopposed estrogen^34^, the estrogen-only versus combined distinction largely reflects this guideline rather than clinical choice, so we modeled estrogen dose regardless of formulation. Exposure was operationalized two ways: a time-varying binary indicator (any systemic HRT versus none) and quarterly oral-equivalent dose categories (none; low, less than 0.5 mg/day; medium, 0.5 to 1.5 mg/day; high, greater than 1.5 mg/day), with bands defined in Appendix C. The binary indicator was derived from the dose categories so both analyses used identical person-time, and a 30-day pharmacological washout defined the end of use. ATC codes, conversion factors, dose-extraction tiers, and quarterly aggregation are detailed in Appendix C.1 through C.8.

### Outcomes

Primary outcomes were sustained EDSS 4.0, sustained EDSS 6.0, and relapse. EDSS 4.0 represents the beginning of ambulatory limitation, while EDSS 6.0 represents the need for unilateral walking assistance^21^. Disability milestones were sustained (i.e., persistence over a subsequent 182-day confirmation window) and were rejected only if EDSS fell below the threshold within that window and the last assessment in the window remained below it. The event date was the date the threshold was first crossed. EDSS assessments up to 182 days beyond study exit were used for confirmation. The full iterative algorithm is provided in Appendix D.1.

Relapses recorded in SMSreg were analyzed as recurring events using the Andersen-Gill extension of Cox regression, which permits multiple events per participant under the time-varying exposure structure.

### Covariates

Baseline covariates were continuous year of entry, educational attainment, country of birth (i.e., Sweden versus abroad), MS phenotype, disease duration from symptom onset to entry, and 8 national regions (i.e., riksområden). DMT class (i.e., none, platform/moderate-efficacy, high-efficacy, anti-CD20; Appendix E) and continuous age were time-varying. For EDSS outcomes the baseline hazard was stratified by baseline EDSS; for relapses it was stratified by the number of relapses in the prior 24 months, with baseline EDSS as a covariate. Full definitions and DMT minimum episode durations are in Appendices E and G. Missing education (45/7,623, 0.59%) and country of birth (<5/7,623) were singly imputed by chained equations (Appendix H).

### Statistical Analysis

Cohort characteristics were described at entry, stratified by any HRT use during follow-up. For sustained EDSS 4.0 and 6.0 we plotted unadjusted Simon-Makuch failure curves by time-varying binary HRT status, an extension of the Kaplan-Meier estimator for a time-varying exposure in which each woman contributes person-time to the current-use curve while using HRT and to the no-use curve otherwise^35^; for recurring relapses every relapse was counted and women remained at risk afterwards, so panel C shows how relapses accumulate over time rather than the probability of a relapse (Figures 2 and S2). We also reported unadjusted incidence rates by time-varying binary HRT status and dose category. Event counts, person-years, and rates for any cell below 5, and for any cell from which a count below 5 could be recovered by subtraction from a reported total, were suppressed in accordance with the ethical approval for this study.

**Figure 2.**
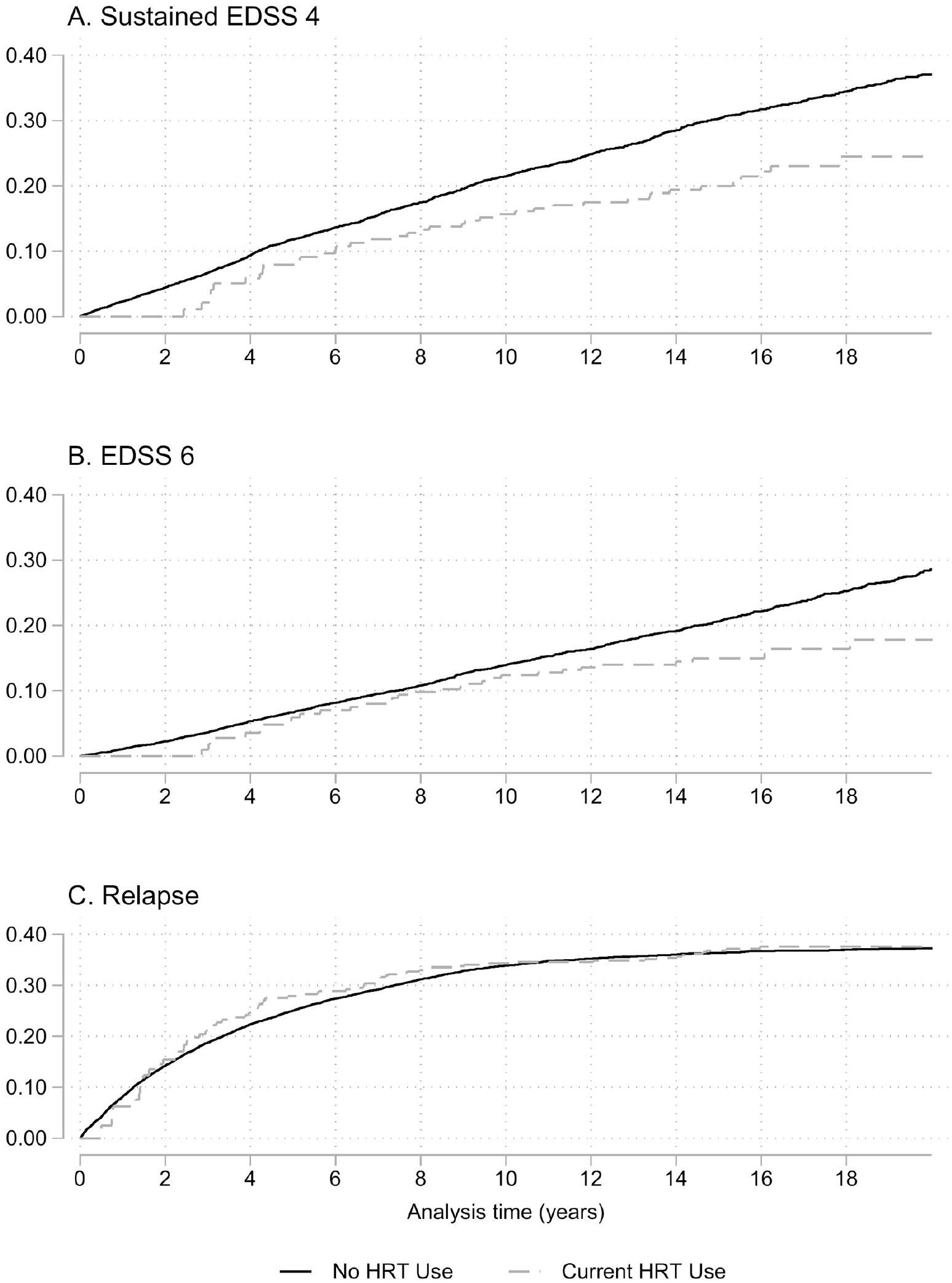
Unadjusted Simon-Makuch failure curves for sustained EDSS 4.0 and 6.0 (A, B) and cumulative recurring relapses (C) by HRT status (primary age 40+ cohort).

We fit Cox proportional-hazards models with time-varying HRT exposure, adjusting for all covariates above, with cluster-robust standard errors for within-person correlation from exposure splitting; separate models were fit for each exposure operationalization and outcome. Recurring relapses used the Andersen-Gill extension. Proportional hazards were assessed with Schoenfeld residual tests.

HRT exposure, DMT class, and age change during follow-up, so HRT was modeled as a time-varying covariate within a Cox framework to align person-time with current exposure status and avoid immortal-time and prevalent-user biases that arise when a time-dependent treatment is treated as fixed at baseline. Time-varying Cox models remove confounding only by the measured, time-updated covariates; they cannot adjust for unmeasured time-varying confounders (e.g., menopausal status and health-seeking behavior) or for confounding affected by prior exposure, and the resulting estimates are therefore conditional associations rather than causal effects.

The binary and dose-category analyses address the same question and were not treated as independent hypotheses; significance was evaluated at alpha = 0.05, and no interaction analyses were prespecified.

### Sensitivity Analyses

Because the full 40-to-60 cohort mixes pre- and post-menopausal women, the unexposed group conflates HRT non-use with retained endogenous estrogen. To address this imprecision, we repeated the primary models in a post-hoc cohort with the entry-age floor raised to 50 years (n = 4,725) to reduce dilution from pre-menopausal unexposed person-time (Appendix I.4).

To assess whether findings were driven by the primary progressive subgroup, we repeated the primary analysis restricted to women with relapsing-remitting MS at diagnosis (Appendix I.6).

All analyses were performed using Stata version 19 (StataCorp, College Station, TX).

### Standard Protocol Approvals, Registrations, and Patient Consents

The Regional Ethical Review Board in Stockholm approved the study (2017/700-31/4), and the Swedish Ethical Review Authority approved an amendment (2021-02384). Individual informed consent was waived because the study used pseudonymized registry data.

### Data Availability

Individual-level data from the Swedish registries cannot be shared publicly due to Swedish law. Researchers may apply for data access through Statistics Sweden (SCB) and the Swedish National Board of Health and Welfare (Socialstyrelsen). Access to the Swedish MS Registry requires a separate application to the registry holder. Analysis code is available from the corresponding author upon reasonable request. No study protocol was registered.

## Results

### Study Population

Of 9,949 women in the Swedish MS Registry aged 40 or older at baseline, alive and resident in Sweden at cohort entry, and with EDSS assessments available, 7,623 aged 40 to 60 at cohort entry met the remaining criteria and formed the analysis cohort (Figure 1). Median follow-up was 13 years (Q1 8, Q3 19). During follow-up, 1,192 (15.6%) initiated systemic HRT. Most participants (94%) had relapsing-remitting MS at diagnosis, and 427 (6%) had primary progressive MS. Baseline EDSS was lower among HRT initiators vs non-initiators (mild EDSS [0 to 3.5]: 84% versus 76%; severe EDSS [6+]: 8% versus 14%). Among HRT initiators, median age at first HRT dispensing was 51.3 years (Q1 48.5, Q3 53.8), consistent with the mean age at natural menopause of 51 years in the Swedish population^36^ and with perimenopausal onset.

HRT initiators were slightly younger at baseline (median 41 versus 42 years; 67% versus 58% in the 40-to-44 band), entered the cohort earlier, and had more relapses in the prior 24 months; phenotype, disease duration, prior hysterectomy, education, parity, and country of birth were similar (Table 1).

**Table 1.** Baseline characteristics at cohort entry, by any HRT use during follow-up (primary age 40+ cohort).

| **No. (Column %) or Median (Q1, Q3)** | **Total**<br>**N=7,623** | **Never**<br>**N=6,431** | **HRT Use**<br>**N=1,192** |
| --- | --- | --- | --- |
| Age | 42 (40, 49) | 42 (40, 50) | 41 (40, 46) |
| Age at baseline |  |  |  |
| 40-44 | 4,551 (60) | 3,748 (58) | 803 (67) |
| 45-49 | 1,293 (17) | 1,053 (16) | 240 (20) |
| 50-54 | 996 (13) | 880 (14) | 116 (10) |
| 55-60 | 783 (10) | 750 (12) | 33 (3) |
| MS Phenotype at Diagnosis |  |  |  |
| Primary Progressive | 427 (6) | 376 (6) | 51 (4) |
| Relapsing-Remitting | 7,196 (94) | 6,055 (94) | 1,141 (96) |
| Disease duration at baseline (years) | 9 (3, 16) | 9 (3, 16) | 8 (2, 15) |
| Highest education level |  |  |  |
| Pre-secondary | 656 (9) | 578 (9) | 78 (7) |
| Secondary | 4,581 (60) | 3,880 (60) | 701 (59) |
| University Degree | 2,386 (31) | 1,973 (31) | 413 (35) |
| Born abroad | 894 (12) | 762 (12) | 132 (11) |
| Total number of births (parity) |  |  |  |
| 0 | 1,688 (22) | 1,473 (23) | 215 (18) |
| 1 | 1,463 (19) | 1,226 (19) | 237 (20) |
| 2 | 3,100 (41) | 2,557 (40) | 543 (46) |
| 3 | 1,065 (14) | 916 (14) | 149 (12) |
| 4+ | 307 (4) | 259 (4) | 48 (4) |
| EDSS Baseline |  |  |  |
| 0-3.5 | 5,908 (78) | 4,912 (76) | 996 (84) |
| 4-5.5 | 703 (9) | 608 (9) | 95 (8) |
| 6+ | 1,012 (13) | 911 (14) | 101 (8) |
| Number of relapses in 24 months before baseline |  |  |  |
| 0 | 5,990 (79) | 5,120 (80) | 870 (73) |
| 1 | 1,223 (16) | 993 (15) | 230 (19) |
| 2 | 318 (4) | 245 (4) | 73 (6) |
| 3+ | 92 (1) | 73 (1) | 19 (2) |
| Residence region |  |  |  |
| Stockholm | 1,868 (25) | 1,544 (24) | 324 (27) |
| Ostra Mellansverige | 1,353 (18) | 1,151 (18) | 202 (17) |
| Smaland med oarna | 460 (6) | 406 (6) | 54 (5) |
| Sydsverige | 979 (13) | 832 (13) | 147 (12) |
| Vastsverige | 1,554 (20) | 1,286 (20) | 268 (22) |
| Norra Mellansverige | 729 (10) | 627 (10) | 102 (9) |
| Mellersta Norrland | 279 (4) | 239 (4) | 40 (3) |
| Ovre Norrland | 401 (5) | 346 (5) | 55 (5) |
| Calendar Period |  |  |  |
| 2006-2009 | 3,481 (46) | 2,947 (46) | 534 (45) |
| 2010-2014 | 1,595 (21) | 1,239 (19) | 356 (30) |
| 2015-2019 | 1,466 (19) | 1,237 (19) | 229 (19) |
| 2020-2024 | 1,081 (14) | 1,008 (16) | 73 (6) |
| Prior hysterectomy without oophorectomy | 128 (2) | 100 (2) | 28 (2) |
| First DMT during study period |  |  |  |
| None | 1,254 (16) | 1,121 (17) | 133 (11) |
| Platform/Moderate | 4,112 (54) | 3,371 (52) | 741 (62) |
| High-efficacy | 799 (10) | 655 (10) | 144 (12) |
| Anti-CD20 | 1,458 (19) | 1,284 (20) | 174 (15) |
| Years of followup | 13 (8, 19) | 13 (7, 19) | 15 (10, 19) |

### Unadjusted Rates

Unadjusted incidence rates were lower during exposed than unexposed person-time for all three outcomes: sustained EDSS 4.0, 15.4 (95% CI 11.2 to 21.2) versus 23.8 (95% CI 22.5 to 25.1) per 1,000 person-years; sustained EDSS 6.0, 10.8 (95% CI 7.6 to 15.4) versus 15.5 (95% CI 14.6 to 16.5); and recurring relapses, 19.1 (95% CI 15.0 to 24.3) versus 31.7 (95% CI 30.6 to 32.9). By dose category, only medium-dose rates could be reported under the registry’s small-cell rules: sustained EDSS 4.0, 14.7 per 1,000 person-years (27 events in 1,837 person-years); sustained EDSS 6.0, 10.0 (21 events in 2,109 person-years); and recurring relapses, 17.3 (43 events in 2,483 person-years). The low- and high-dose cells were suppressed. Full rates, person-years, and event counts appear in Table 2.

**Table 2.** HRT events, rates, and adjusted hazard ratios (primary age 40+ cohort).

| Exposure | Sustained EDSS 4 |  |  |  |  | Sustained EDSS 6 |  |  |  |  | Recurring Relapse |  |  |  |  |
| --- | --- | --- | --- | --- | --- | --- | --- | --- | --- | --- | --- | --- | --- | --- | --- |
|  | Events | Person-Years (PY) | Per 1,000 PY (95% CI) | aHR (95% CI) | p-value | Events | Person-Years (PY) | Per 1,000 PY (95% CI) | aHR (95% CI) | p-value | Events | Person-Years (PY) | Per 1,000 PY (95% CI) | aHR (95% CI) | p-value |
| Binary HRT |  |  |  |  |  |  |  |  |  |  |  |  |  |  |  |
| None | 1,363 | 57,334 | 23.8 (22.5, 25.1) | Reference |  | 1,092 | 70,252 | 15.5 (14.6, 16.5) | Reference |  | 2,967 | 93,488 | 31.7 (30.6, 32.9) | Reference |  |
| Current HRT use | 38 | 2,462 | 15.4 (11.2, 21.2) | 0.70 (0.51, 0.96) | 0.028 | 31 | 2,857 | 10.8 (7.6, 15.4) | 0.80 (0.55, 1.16) | 0.24 | 67 | 3,507 | 19.1 (15.0, 24.3) | 1.00 (0.74, 1.36) | 0.98 |
| Estrogen Dose Category |  |  |  |  |  |  |  |  |  |  |  |  |  |  |  |
| None | 1,363 | 57,334 | 23.8 (22.5, 25.1) | Reference |  | 1,092 | 70,252 | 15.5 (14.6, 16.5) | Reference |  | 2,967 | 93,488 | 31.7 (30.6, 32.9) | Reference |  |
| Low (<0.5mg) | – | – | – | 0.97 (0.33, 2.86) | 0.96 | – | – | – | 1.38 (0.47, 4.09) | 0.56 | – | – | – | 0.73 (0.26, 2.07) | 0.55 |
| Medium (0.5-1.5mg) | 27 | 1,837 | 14.7 (10.1, 21.4) | 0.69 (0.47, 0.99) | 0.046 | 21 | 2,109 | 10.0 (6.5, 15.3) | 0.80 (0.52, 1.23) | 0.31 | 43 | 2,483 | 17.3 (12.8, 23.3) | 0.96 (0.66, 1.39) | 0.83 |
| High (>1.5mg) | – | – | – | 0.68 (0.34, 1.35) | 0.27 | – | – | – | 0.63 (0.28, 1.43) | 0.27 | – | – | – | 1.17 (0.72, 1.92) | 0.52 |

Note: HRT, Hormone Replacement Therapy; aHR, adjusted hazard ratio; CI, confidence interval. All models adjusted for age, calendar year, education, foreign country of birth, MS phenotype, disease duration, region of residence, and time-varying DMT class. Relapse model adjusts additionally for baseline EDSS. EDSS outcomes stratify baseline hazards by baseline EDSS. Relapse model stratifies baseline hazards by number of relapses in 24 months prior to baseline. Model standard errors clustered by patient id. En-dashed cells (–) are suppressed to comply with registry privacy restrictions because the underlying event count falls below the registry's disclosure threshold; within the estrogen-dose categories a further cell is suppressed where necessary to prevent a suppressed count from being recovered by subtraction from the published total.

### Primary Analyses

After multivariable adjustment, time-varying HRT use (any versus none) was associated with a lower hazard of sustained EDSS 4.0 (adjusted hazard ratio [aHR] 0.70, 95% CI 0.51 to 0.96; p = 0.028) and was not associated with sustained EDSS 6.0 (aHR 0.80, 95% CI 0.55 to 1.16; p = 0.24) or recurring relapses (aHR 1.00, 95% CI 0.74 to 1.36; p = 0.98) (Table 2; full models in Table S3).

Dose-category analyses showed no consistent association between HRT exposure and EDSS progression or recurring relapses. Estimates were imprecise; the only confidence interval in the primary cohort that excluded unity was for medium-dose exposure and sustained EDSS 4.0 (aHR 0.69, 95% CI 0.47 to 0.99), and no consistent dose-response pattern was observed (Tables 2 and S4).

### Proportional Hazards Assessment

The global Schoenfeld test did not reject proportional hazards for the EDSS 4.0 (p = 0.09) or EDSS 6.0 (p = 0.78) models. For recurring relapses the global test was significant (p < 0.001), driven by calendar year, age, baseline EDSS, and region rather than HRT. The HRT term showed no departure from proportionality in any of the three binary-exposure models (EDSS 4.0 p = 0.65; EDSS 6.0 p = 0.22; relapse p = 0.31), so the Cox specification was retained (Appendix I.2).

### Covariate Associations

Across all models, primary progressive phenotype at diagnosis strongly predicted disability milestones (aHR 1.94, 95% CI 1.56 to 2.40 for EDSS 4.0; aHR 1.73, 95% CI 1.40 to 2.13 for EDSS 6.0). Anti-CD20 therapy showed the largest reduction in relapse risk (aHR 0.46, 95% CI 0.38 to 0.56) but, consistent with confounding by indication, higher disability rates (EDSS 4.0 aHR 1.46, 95% CI 1.22 to 1.74), as did high-efficacy non-anti-CD20 therapy (EDSS 4.0 aHR 1.37, 95% CI 1.09 to 1.73). Foreign birth predicted higher EDSS 4.0 risk (aHR 1.37, 95% CI 1.16 to 1.62) and university education lower risk (aHR 0.75, 95% CI 0.66 to 0.86). Regional variation followed no consistent direction; full results appear in Tables S3 and S4.

### Sensitivity Analyses

In the entry-age-50 sensitivity cohort (n = 4,725; 659 HRT initiators, 13.9%), estimates tracked the primary analysis (Tables S1, S2). The sustained EDSS 4.0 association was similar and reached significance (aHR 0.62, 95% CI 0.40 to 0.95; p = 0.029), while sustained EDSS 6.0 (aHR 0.90, 95% CI 0.57 to 1.42) and recurring relapses (aHR 0.95, 95% CI 0.60 to 1.52) remained near null. Dose-category estimates mirrored the primary pattern (medium-dose EDSS 4.0 aHR 0.55, 95% CI 0.33 to 0.94; Tables S5, S6). Median age at first dispensing in this cohort was 53.2 years (Q1 51.6, Q3 55.3).

Restricting the age-40+ cohort to relapsing-remitting MS reproduced the full-cohort results (EDSS 4.0 aHR 0.71, 95% CI 0.51 to 0.97; EDSS 6.0 aHR 0.78, 95% CI 0.53 to 1.16; relapses aHR 1.02, 95% CI 0.75 to 1.38; Table S7), as did the relapsing-remitting entry-age-50 cohort (Table S8).

## Discussion

In a nationwide cohort of 7,623 Swedish women with MS followed through the modern high-efficacy DMT era, current systemic HRT use was associated with a lower hazard of sustained EDSS 4.0, and no association with sustained EDSS 6.0 or recurring relapses. Dose-category point estimates for EDSS 4.0 were below 1.0 at every dose level, with wide confidence intervals. In light of the clinical debate articulated by Voskuhl and Magyari regarding whether menopause-related hormone loss contributes to disability accrual in MS, these findings do not provide clear evidence that HRT exerts disease-modifying effects comparable to established DMTs. Rather, the observed association with EDSS 4.0 should be interpreted cautiously and warrants replication in cohorts with verified menopausal status and refined exposure classification before stronger conclusions can be drawn.

The only prior population-based cohort study, from Denmark, reported an increased relapse risk with HRT that the authors attributed to 1996 to 2005^16^. A small (n=16) prospective study in Finland found no adverse changes in progression-related biomarkers over 12 months of menopausal hormone therapy^37^. A phase 2 randomized trial showed that oral estriol added to glatiramer acetate reduced relapses at the pre-specified significance level but was not designed to assess disability^38^. We found no evidence of an increased relapse risk, consistent with the Danish estimates outside their earliest calendar period. The association with a lower risk of sustained EDSS 4.0 has not been previously reported in population-based data.

Both Kopp et al.^16^ and the present study modeled HRT as a time-dependent exposure, but the analyses differ in ways that bear on the relapse comparison. Kopp et al. included women with HRT dispensed before cohort entry, whereas we applied a new-user design; they carried exposure forward for four months after a prescription was expected to run out, whereas we used a 30-day washout; they characterized exposure by cumulative duration and regimen, whereas we converted dispensings to an oral estradiol-equivalent daily dose with quarterly aggregation; and their cohort comprised DMT-treated women with relapsing-remitting MS aged 40 to 79 followed from 1996, whereas ours spans 2006 to 2025 with anti-CD20 therapy as a distinct time-varying class. The excess relapse risk they observed was driven by the 1996 to 2005 period, which precedes our study window, so their findings and ours are consistent rather than contradictory.

Our results separate the disability and relapse domains in the same direction reported after natural menopause, where relapse rates decline while disability accumulation accelerates^8,11^. If exogenous estrogen acts mainly through neuroprotective rather than anti-inflammatory pathways^4,5^, effects on disability milestones rather than relapse frequency would be expected, matching the point estimates. The dose categories do not establish a dose-response relationship: EDSS 4.0 estimates were below 1.0 at every dose level, but the low- and high-dose event counts and rates could not be reported under the registry’s small-cell rules, confidence intervals were wide at every level and crossed unity at every level except medium dose for EDSS 4.0 in the primary cohort, and no monotonic gradient was present.

The contrast between EDSS 4.0 and EDSS 6.0 likely reflects both biological stage-dependence and non-causal selection. Biologically, EDSS 4.0 may mark a phase where neuroaxonal reserve and functional compensation remain sufficient for modest neuroprotective effects of estrogen to delay sustained disability, whereas EDSS 6.0 reflects more advanced structural injury less amenable to hormonal modulation, consistent with frameworks in which estrogen supports neuroprotection rather than late-stage disease modification.

A non-causal explanation cannot be excluded. Women initiating HRT had lower baseline EDSS than non-initiators, consistent with a healthy-user profile (e.g., better functional status, greater functional engagement, greater physiological reserve). Stratifying the baseline hazard by baseline EDSS does not capture these differences, so a lower hazard of sustained EDSS 4.0 may reflect residual baseline function rather than an effect of HRT, preferentially delaying a mobility-based milestone like EDSS 4.0 without altering progression to EDSS 6.0. Power was also lower for EDSS 6.0. These mechanisms cannot be disentangled here.

Multiple cohorts report accelerated disability after menopause^8–10,12^, making a lower disability hazard among women dispensed HRT biologically plausible. The unexposed group in the 40- to-60 cohort still includes pre-menopausal women, diluting the contrast toward the null; the entry-age-50 and relapsing-remitting sensitivity analyses, which reduce this dilution, yielded EDSS 4.0 estimates of similar magnitude (i.e., aHR 0.62 and 0.71, respectively). The finding is therefore not an artifact of the primary progressive subgroup, but without replication in cohorts with verified menopausal status the EDSS 4.0 signal should not be over-interpreted.

Two features of the exposure distribution caution against over-interpretation. HRT initiators entered during earlier calendar periods (Table 1), corresponding to an earlier DMT era and less frequent EDSS measurement; although we adjusted for year of entry, residual calendar-related confounding cannot be excluded. Estrogen-only and combined preparations were also collapsed, because Swedish guidelines mandate progestogen for women with an intact uterus^34^; progestogen type may have differential neurological effects^39^, but the low progestogen doses and few exposed events precluded examining formulation as a modifier.

This analysis has several limitations. The Prescribed Drug Register captures dispensing rather than consumption, and the bridging and quarterly aggregation rules add measurement imprecision that would generally bias toward the null. Menopausal status was not recorded in any registry, so the comparison is between women with and without dispensed HRT rather than estrogen-depleted versus estrogen-replete women, conflating HRT use with menopausal timing and symptom-driven treatment-seeking; the entry-age-50 analysis reduces but does not eliminate this confounding. Beyond stroke and venous thromboembolism, other cardiovascular comorbidity was not excluded, and because HRT prescribing tracks cardiovascular risk, residual confounding by cardiovascular health cannot be excluded.

Exposed person-time was limited, constraining dose-specific precision. Inverse-probability-weighted marginal structural models were attempted but abandoned because covariate imbalances could not be resolved. If women with more active disease are less likely to initiate HRT, the EDSS 4.0 association could partly reflect this residual confounding.

In conclusion, in this large contemporary cohort systemic HRT use showed no indication of increased relapse risk or accelerated severe disability progression. For women with MS considering HRT for menopausal symptoms, these data do not support concerns about worsened disease activity in the high-efficacy DMT era. The lower hazard of sustained EDSS 4.0 should be read as a signal, not evidence of a disease-modifying benefit, because it was not mirrored at EDSS 6.0, the dose categories showed no consistent gradient, and the baseline EDSS imbalance is consistent with residual healthy-user bias. These findings derive from a predominantly white Nordic population with high anti-CD20 uptake and may not generalize to other settings. Replication in cohorts with verified menopausal status is needed for a clearer interpretation of the present study’s findings.

## Supporting information

All supplemenetary material including RECORD checklist

## Author Contributions

TPC: Conceptualization, Methodology, Software, Formal analysis, Data curation, Writing – original draft. KAM: Conceptualization, Methodology, Supervision, Writing – review and editing. AP: Methodology, Writing – review and editing. RHO: Methodology, Writing – review and editing. KF: Conceptualization, Data curation, Supervision, Writing – review and editing.

## Acknowledgements

We are grateful for the support and data contributions from women with MS and their clinicians who facilitate the MS Registry. We also thank Thomas Frisell for his support. Claude (Anthropic) and GPT (OpenAI) reviewed the Stata code and drafted appendices from it. TPC verified their corrections and the appendices.

## Supplementary Material

See Technical Appendix for detailed methods documentation.

## Supplementary Tables

- Table S1: Baseline characteristics by HRT use among the entry-age-50 sensitivity cohort
- Table S2: HRT events, rates, and adjusted hazard ratios among the entry-age-50 sensitivity cohort
- Table S3: Complete adjusted model results for binary HRT exposure (primary age 40+ cohort)
- Table S4: Complete adjusted model results for estrogen dose categories (primary age 40+ cohort)
- Table S5: Complete adjusted model results for binary HRT exposure among the entry-age-50 sensitivity cohort
- Table S6: Complete adjusted model results for estrogen dose categories among the entry-age-50 sensitivity cohort
- Table S7: HRT events, rates, and adjusted hazard ratios among relapsing-remitting MS subgroup (primary age 40+ cohort)
- Table S8: HRT events, rates, and adjusted hazard ratios among relapsing-remitting MS subgroup (entry-age-50 sensitivity cohort)

## Figures

- Figure S1: Sequential exclusion flowchart, starting from women alive and resident in Sweden at cohort entry (entry-age-50 sensitivity cohort)
- Figure S2: Unadjusted Simon-Makuch failure curves for sustained EDSS 4.0 and 6.0 (A, B) and cumulative recurring relapses (C) by HRT status (entry-age-50 sensitivity cohort)

