## Supplementary material for "Hormone Replacement Therapy and Disability Progression in Women with Multiple Sclerosis": All supplemenetary material including RECORD checklist

### Table of Contents


- Appendix A. Data Sources and Linkage
- Appendix B. Cohort Construction
- Appendix C. HRT Exposure Definition
- Appendix D. Outcome Definitions
- Appendix E. DMT Exposure Definition
- Appendix F. Time-Varying Covariate Integration
- Appendix G. Baseline Covariate Specifications
- Appendix H. Missing Data Handling
- Appendix I. Statistical Analysis
- References
- Supplementary Tables S1 to S8 (after the appendix references)
- Supplementary Figures S1 and S2
- RECORD checklist


### Appendix A. Data Sources and Linkage

*A.1 Registry Components*

| Registry | Coverage | Variables Used |
|---|---|---|
| Swedish MS Registry (SMSreg) | Specialty MS care, clinician-entered | EDSS scores with dates, MS phenotype, relapse dates, DMT start/stop with ongoing-treatment flag, demographics |
| Prescribed Drug Register (PDR) | July 2005 onwards, all dispensed prescriptions | ATC code, dispensing/prescribing dates, package size, DDD per package, packages, strength, formulation |
| National Patient Register (NPR) | Inpatient 1987+; specialist outpatient 2001+ | Up to 30 ICD-10 diagnoses and 30 procedure codes per visit, with dates |
| Swedish Cancer Registry | All malignancies, >96% ascertainment | ICD-O-3 topography/morphology, cancer diagnosis date |
| LISA (Statistics Sweden) | Annual, longitudinal | Education, income, employment |
| Register of the Total Population (RTB) | Annual | Municipality/county of residence |
| Medical Birth Registry | 1973 onwards | Birth dates of biological children, parity |

| Registry | Coverage | Variables Used |
|---|---|---|
| Migration Register | All-time | Immigration/emigration dates |
| Cause of Death Register | All-time | Date and causes of death |

The PDR does not capture in-hospital medications or dispensings before July 2005; NPR diagnoses before 2001 are restricted to inpatient encounters.

*A.2 Linkage and Reporting*

All Swedish residents receive a unique personal identification number. The National Board of Health and Welfare and Statistics Sweden performed deterministic linkage on these identifiers and pseudonymized the data with project-specific study IDs before delivery. The study follows the RECORD extension of STROBE. Fulfillment of the McDonald diagnostic criteria is a mandatory SMSreg field, so case ascertainment reflects neurologist-recorded MS diagnoses under the contemporaneous criteria. Individual-level data cannot be shared publicly under Swedish law; access requires application to Statistics Sweden and the National Board of Health and Welfare, with a separate application to the SMSreg holder.

### Appendix B. Cohort Construction

This appendix documents how the analysis cohort is assembled: the source population and sequential selection steps (B.1); the computation of study entry, baseline EDSS, and migration history (B.2); and the inclusion (B.3) and exclusion (B.4) criteria in RECORD checklist form. The selection steps in B.1 and the criteria in B.3 and B.4 describe the same cohort from procedural and declarative perspectives.

*B.1 Source Population and Restrictions*

The source population is every individual in SMSreg. Selection proceeds in sequential steps before study entry is computed, followed by a final age check.

| Step | Restriction |
|---|---|
| 1 | Restrict to female sex |
| 2 | Drop if age would fall outside 40-60 across the study window |
| 3 | Drop if MS phenotype, first DMT start date, date of birth, or pre-2006 last registry activity is missing. If MS diagnosis date is missing, use first DMT start as a proxy; drop if both missing or post-2024 |

| Step | Restriction |
|---|---|
| 4 | Drop if only one EDSS assessment exists, or if the last EDSS precedes study entry |
| 5 | Drop if not resident in Sweden at entry (B.2), or if death or emigration falls on or before study entry |
| 6 | After entry is computed, drop if age at study entry exceeds 60 |

DMT engagement (Step 3) confirms an actionable clinical diagnosis; the first-DMT-start proxy for a missing diagnosis date is rarely needed and is documented because the manuscript Methods refer to it.

*B.2 Study Entry, Baseline EDSS, and Migration*

Study entry is the latest of the participant's 40th-birthday anniversary (50th for the sensitivity cohort), the MS diagnosis date (or first-DMT proxy), and 1 January 2006. Age at entry is computed as completed years to the birthday anniversary rather than `floor(days/365.25)`, preventing leap-year drift across age bands. Entry is limited to 31 December 2024 to ensure at least one year of potential follow-up.

Baseline EDSS is the most recent assessment before study entry; when none exists, the earliest recorded EDSS is used. When multiple scores share a date, the maximum is retained (conservative for progression outcomes).

Migration history is reconstructed from all dated immigration/emigration events. A participant is excluded if her last emigration occurred before entry without return, or if a post-entry immigration has no prior post-entry emigration (non-residence at entry). For those retained, the first post-entry emigration is the censoring date.

*B.3 Inclusion Criteria (all required)*

1. Female sex
2. MS diagnosis recorded in SMSreg, with a recorded phenotype and diagnosis date
3. Age 40-60 at entry (50-60 in the sensitivity cohort)
4. At least two EDSS assessments, the last at or after entry
5. At least one DMT record
6. Study entry 1 January 2006 to 31 December 2024
7. Alive and resident in Sweden at entry (death or emigration on or before entry excludes)

*B.4 Exclusions*

| Exclusion | Definition | Rationale |
|---|---|---|
| Prior surgical menopause | Bilateral oophorectomy on or before entry (procedure codes LAE2, LAF1, LAF3) | Differs mechanistically from natural menopause |
| Malignant cancer | Any malignant neoplasm before entry (ICD-10 C00-C97 incl. C77-C80; excluding C44); earliest date across Cancer Registry and NPR | May contraindicate HRT; treatment may alter MS course |
| Venous thromboembolism | ICD-10 I26, I80, I82 on or before entry | Absolute contraindication to systemic estrogen |
| Stroke | ICD-10 I60, I61, I63, I64 on or before entry | Cerebrovascular disease may contraindicate HRT |
| Prior intensive immunosuppression | Mitoxantrone (SMSreg) or HSCT (NPR codes QA080, QA100) before entry | Fundamentally alters disease course |
| Birth during follow-up | Live birth after entry; censored at estimated conception (270 days before birth); excluded if conception falls on or before entry | Pregnancy alters hormonal milieu and MS course |
| Prior HRT use | Any systemic HRT dispensing before entry | New-user design avoids prevalent-user bias |

**At-risk restriction (not a cohort exclusion):** Women with baseline EDSS >= 4.0 are removed from the at-risk population for the sustained-EDSS-4.0 outcome, and likewise >= 6.0 for EDSS 6.0. The recurring-relapse analysis uses the full cohort without a baseline-EDSS restriction.

**Left-censoring of HRT history:** Because the PDR begins July 2005, women entering in 2006 have at most six months of observable pre-entry history, so some pre-July-2005 HRT users may be misclassified as new users (most likely among early entrants). The new-user design is therefore approximate.

### Appendix C. HRT Exposure Definition

*C.1 Relevant ATC Codes*

**Estrogen-only systemic:** G03CA03 (estradiol, oral/transdermal), G03CA04 (estriol, oral), G03CA53 (estradiol combinations), G03CA57 (conjugated estrogens, oral).

**Combined estrogen-progestogen, continuous:** G03FA01, G03FA12, G03FA14, G03FA15, G03FA17.

**Combined estrogen-progestogen, sequential:** G03FB01, G03FB05, G03FB06, G03FB08, G03FB09, G03FB11.

**Tibolone:** G03CX01 (oral-equivalent assigned via conversion, Appendix C.5).

**Excluded:** G03HB01 (cyproterone + ethinylestradiol; anti-androgen/contraceptive). Standalone progestogens (G03DA02, G03DC02, G03DC03, G02BA03) do not define systemic HRT. Swedish guidelines require combined HRT for women with an intact uterus, so the estrogen-only versus combined distinction largely reflects guideline-mandated progestogen add-on; all systemic dispensings contribute estrogen dose regardless of progestogen co-dispensing.

Vaginal preparations are removed from the systemic classification when the formulation field (`form`) contains "Vaginal" or "Vagitorium". A separate route-classification step additionally labels a dispensing as vaginal when `form` or `nameform` contains "Vagi", but route labeling does not alter systemic inclusion.

*C.2 Days Supply*

Days supply per dispensing is defined daily doses per package times number of packages, rounded up. Negative package counts are converted to absolute values. One product-specific fallback supplies a missing `packddd` (G03FA15, package "3 x 28 styck" defaults to 84 DDD). Dispensings with missing days-supply inputs are logged by ATC before dose extraction.

*C.3 Exposure Period Construction*

Dose-category exposure periods from quarterly aggregation (Appendix C.7) are consolidated into continuous intervals using a 90-day merge window and a 30-day pharmacological washout (effective grace period up to 120 days). Consecutive intervals merge when the gap is <= 90 days and the dose category matches; larger gaps or category transitions start a new period. The 90-day window approximates one refill cycle; the 30-day washout reflects pharmacological carryover.

**Unified construction:** Both the binary indicator (Approach 1) and the dose-category variable (Approach 2) are built from a single exposure dataset; the binary indicator is defined as "any positive dose category," guaranteeing identical person-time across analyses. Dispensings with failed dose extraction (zero/missing total oral-equivalent) are excluded from analyzable intervals; the descriptive `hrt_status` flag and the prevalent-user exclusion are generated before those rows are dropped.

*C.4 Estrogen Dose Extraction (3-Tier Method)*

Per-unit estrogen content (mg) is extracted in three tiers, in order of preference:

- **Tier 1, structured strength field** (standalone products G03CA03, G03CA04, G03CA53, G03CX01): the numeric strength field reports estrogen content; microgram encodings (`mcg`, `mikrogram`, `mikrog/24 timmar`, `mikrog/dos`, `µg`) are rescaled to mg.
- **Tier 2, parsed strength text** (combined continuous G03FA01, G03FA12, G03FA14, G03FA15, G03FA17, and the sequential dydrogesterone combination G03FB08): strength text gives both components as `"X mg/Y mg"` (oral), `"X mg + X mg/Y mg"` (sequential G03FB08), or transdermal microgram form; estrogen is the first component. Swedish decimal commas are converted to dots before regex extraction.
- **Tier 3, product nameform lookup** (sequential combined products lacking structured/parseable strength): per-tablet doses assigned from FASS/EMC/MPA Sweden references.

| Product | ATC | Estrogen per unit | Route |
|---|---|---|---|
| Trisekvens | G03FB05 | 1.786 mg/tablet | Oral |
| Femasekvens | G03FB05 | 2.0 mg/tablet | Oral |
| Novofem | G03FB05 | 1.0 mg/tablet | Oral |
| Sequidot | G03FB05 | 0.05 mg/24h | Transdermal |
| Estalis Sekvens | G03FB05 | 0.05 mg/24h | Transdermal |
| Trivina | G03FB06 | 1.846 mg/tablet | Oral |
| Divina Plus | G03FB06 | 2.0 mg/tablet | Oral |
| Premelle Sekvens | G03FB06 | 0.625 mg/tablet | Oral |
| Femoston (fallback only; Tier 2 reads 1 or 2 mg from the strength text) | G03FB08 | 2.0 mg/tablet | Oral |
| Cyclabil | G03FB09 | 2.0 mg/tablet | Oral |
| Lenzetto | G03CA03 | 0.05 mg equiv/spray | Spray |
| Femanor | G03FA01 | 1.0 mg/tablet | Oral |
| Activelle (generic) | G03FA01 | 1.0 mg/tablet | Oral |
| Premelle (continuous) | G03FA12 | 0.625 mg/tablet | Oral |
| Indivina | G03FA12 | 1.0 mg/tablet | Oral |
| Angemin | G03FA17 | 1.0 mg/tablet | Oral |

*C.5 Oral-Equivalent Conversion*

All doses are converted to an oral estradiol-equivalent scale so average daily dose (mg/day) is comparable across routes. Clinical-equivalence factors[1,2] are used rather than WHO DDD ratios[3], which would impose clinically nonsensical route asymmetries.

| **Route/Product** | **Factor** | **Anchor** | **Source** |
|---|---|---|---|
| Oral estradiol (G03CA03, G03CA53 oral) | x1 | 1 mg/day oral 17-beta-estradiol = standard dose | [1,2] |
| Transdermal patch (G03CA03 transdermal; G03FB05 Sequidot/Estalis) | x20 | 50 mcg/day systemic = 1 mg/day oral | [4,5] |
| Transdermal spray (Lenzetto) | x20 on 0.05 mg/spray placeholder | Coded as systemic equivalent of a 50 mcg/day patch | [5] |
| Estradiol gel (G03CA53 gel) | x1 | 1 mg/day applied gel = standard dose | [2] |
| Conjugated equine estrogens (G03CA57; Premelle G03FA12, G03FB06) | x1.6 | 0.625 mg/day CEE = 1 mg/day oral E2 | [1,6] |
| Estriol, oral (G03CA04) | x0.1 | Weak ER-alpha affinity, rapid clearance | [2,7] |
| Tibolone (G03CX01) | x0.1 | Pragmatic dose-categorization factor | [8] |

**Notes.** The transdermal x20 factor (not x40) follows clinical dose equivalence: 50 mcg/day transdermal, 1 mg/day oral estradiol, and 0.625 mg/day CEE are treated as equipotent standard doses[1,2,5]. The tibolone and oral-estriol x0.1 factors are pragmatic dose-categorization conventions (placing both in the “Low” category), not pharmacokinetic equivalences[7,8]; sensitivity analyses excluding tibolone are supported by the data structure. Estradiol-valerate products (Trivina, Cyclabil, Divina Plus, Indivina) are entered at face-value milligrams consistent with NAMS/FASS/SmPC convention[1,9]; because EV (MW 356.5) releases only the estradiol base (MW 272.4)[10,11], this introduces a ~24% systematic overestimate of free-E2 content for EV products, a known limitation retained for comparability with the published literature.

*C.6 Total Oral-Equivalent Dose per Dispensing*

For **oral** products, total oral-equivalent is per-tablet content x tablets dispensed. For **non-oral** products (patch, gel, spray), `oral_equiv_mg` is a daily oral-equivalent rate, so total dose is rate x days supply. Splitting by route avoids multiplying a daily rate by a unit count, which would underestimate dose for products applied every 3-4 days.

*C.7 Quarterly Dose Aggregation*

Estrogen dose is aggregated to person-specific 91-day quarters anchored to each individual's first HRT dispensing. Each dispensing is mapped to the quarter(s) it overlaps; the oral-equivalent mg contributed is the daily rate times overlap days. Within each quarter, total dispensed oral-equivalent is divided by days actually covered (`q_days_covered`), so partial-quarter dispensings yield dose estimates representative of treatment days. This first-dispensing-anchored quarterly panel feeds both the dose-category and the derived binary exposure.

*C.8 Exposure Approach Summary*

| **Approach** | **Variable** | **Type** | **Description** |
|---|---|---|---|
| 1 | hrt_tv | Binary (0/1) | Current HRT use: any vs none (positive dose category) |
| 2 | hrt_dosecat | Categorical (0-3) | None / Low / Medium / High |

| **Category** | **Oral-equivalent dose** | **Interpretation** |
|---|---|---|
| 0: None | 0 mg/day | No current HRT |
| 1: Low | >0 to <0.5 mg/day | Half-dose products, estriol, tibolone |
| 2: Medium | 0.5 to 1.5 mg/day | Standard 1 mg oral estradiol |
| 3: High | >1.5 mg/day | 2 mg oral, high-dose transdermal |

Categories are built from explicit non-overlapping bins; a post-construction assertion verifies every non-missing daily dose receives a category.

*C.9 Key References for Conversion Factors*

The oral-equivalent conversion factors in Appendix C.5 rest on the following sources. Route-specific pharmacokinetics and the oral-versus-transdermal equivalence are taken from Kuhl[2], Balfour and Heel[4], and Goodman[5]. The conjugated-equine-estrogen anchor follows Bhavnani and Stanczyk[6] and the NAMS 2022 position statement[1]. The estriol factor reflects the relative estrogen-receptor binding affinities reported by Kuiper et al.[7], and the tibolone factor follows the International Tibolone Consensus Group[8]. WHO defined daily doses[3] were reviewed but not used as conversion factors. Per-tablet doses for Tier-3 products were taken from the corresponding FASS[9], EMC[12], and Swedish Medical Products Agency[13] summaries of product characteristics; molecular weights for estradiol valerate and estradiol are from PubChem[10,11].

### Appendix D. Outcome Definitions

The three primary outcomes use two frameworks. EDSS milestones (sustained 4.0 and 6.0) are time-to-first-event: follow-up ends at the first confirmed crossing or censoring. Relapses are recurring events analyzed with the Andersen-Gill extension of Cox regression: follow-up continues regardless of prior events. The person-level censoring date (Appendix D.3) is identical across outcomes.

*D.1 Confirmed EDSS Milestone Algorithm*

For each threshold (4.0, 6.0), the candidate milestone date is the earliest EDSS at or above the threshold; the following 182-day window is then assessed. A candidate crossing is **rejected** only when *both* (a) some EDSS within the window falls below the threshold and (b) the last EDSS within the window is below the threshold. On rejection, the crossing assessment is replaced by the last within-window value and the search repeats for the next earliest crossing. If no EDSS occurs within the window, or a transient dip is followed by a final within-window EDSS back at or above threshold, the candidate is retained. Iteration continues until no candidate is rejected; the event date is the earliest remaining EDSS at or above threshold.

**Post-exit confirmation.** EDSS assessments up to 182 days beyond study exit are used for confirmation, so events near study end may be confirmed using post-exit data. The event date is the first-crossing date, not the confirmation date.

*D.2 Recurring Relapse Outcome*

All relapses after entry and on or before exit are retained, with no fixed cap, after dropping duplicate relapse dates within a person; the number of relapse variables is set by the observed maximum. These are analyzed with the Andersen-Gill extension, allowing multiple events per person while maintaining time-varying exposure.

*D.3 Censoring Events*

The person-level exit date is the earliest of: first bilateral oophorectomy, first VTE, first stroke, death, first post-entry emigration, estimated conception (first post-entry birth minus 270 days), the 80th birthday, or administrative end (31 December 2025). Outcomes are not incorporated into the exit date; they are assigned afterward through the time-varying event datasets. The conception-based censor is folded in at baseline-dataset construction so every downstream dataset inherits it; women whose conception censor precedes entry are excluded.

**Appendix E. DMT Exposure Definition**

*E.1 DMT Classification*

| Category | Label | Drugs |
|---|---|---|
| 0 | No DMT | Periods with no active DMT exposure |
| 1 | Platform / moderate-efficacy | Interferon-beta (Avonex, Betaferon, Extavia, Plegridy, Rebif), glatiramer acetate, teriflunomide, dimethyl fumarate |
| 2 | High-efficacy, non-anti-CD20 | Fingolimod, siponimod, ozanimod, ponesimod, cladribine, alemtuzumab, natalizumab |
| 3 | Anti-CD20 monoclonal antibodies | Rituximab (mostly off-label in Sweden), ocrelizumab, ofatumumab |

*E.2 Episode Construction*

SMSreg provides treatment start/stop dates; missing stops are resolved hierarchically: ongoing-flagged episodes extend to study end (2025-12-31); otherwise the episode closes the day before the next start for that person; otherwise it extends to study end. Episode starts before entry are truncated to entry for the first episode. Treating missing stops as ongoing may overestimate DMT duration but preserves exposure continuity.

*E.3 Minimum Episode Durations*

The registry sometimes records same-day start/stop episodes (data-entry artifacts). Pharmacology-based minimum durations prevent spurious treatment gaps:

| Drug | Minimum (days) |
|---|---|
| Avonex, Betaferon, Copaxone, Extavia, Rebif | 7 |
| Plegridy | 14 |
| Aubagio, Dimetylfumarat, Fingolimod, Kesimpta, Mayzent, Ponvory, Skilarence, Vumerity, Zeposia | 30 |
| Natalizumab | 42 |
| Mitoxantrone | 90 |
| Ocrevus, Rituximab, Arzerra IV | 180 |
| Lemtrada, Mavenclad | 365 |

If extending an episode to its minimum would overlap the next start, the stop is moved to the day before that start.

*E.4 Merging and Exclusions*

A 120-day grace period merges consecutive episodes of the same class. Explicit "no treatment" rows are dropped and reconstructed as gaps between active periods. Anti-CD20 episodes whose recorded stop precedes start + 180 days are extended to start + 180 days (B-cell depletion persists beyond the dosing interval), with any subsequent DMT start pushed past the extended stop. Episodes with categories 5-8 or missing (HSCT, experimental, unclassifiable) are dropped, as are same-day stop-reason-2 (patient decision) and stop-reason-99 (administrative) artifacts.

### Appendix F. Time-Varying Covariate Integration

Age is modeled as a continuous time-varying covariate (`age_tv`) advancing at each birthday between entry and exit. HRT, DMT, and age time-varying datasets are combined by sequential pairwise interval intersection within person (overlap start = later start, overlap stop = earlier stop; zero/negative-duration intervals dropped), so each row is a period of constant HRT, DMT, and age status. Because the binary and dose-category exposures originate from the same construction (Appendix C.8), person-time denominators are identical across approaches. A per-person `tvdiagnose` check reports any gaps or overlaps in the merged intervals. Zero-duration first intervals receive a 0.5-day offset; overlapping quarter boundaries are deconflicted by moving the later start to the previous stop plus one day. Each dataset is merged with outcome dates so the event indicator falls in the interval containing the event; persons without an event are censored at the last interval.

**Appendix G. Baseline Covariate Specifications**

*G.1 Continuous Variables*

| Variable | Calculation |
|---|---|
| Age at entry | Completed years from date of birth to study entry by the birthday-anniversary method (Appendix B.2), not floor(days / 365.25) |
| Disease duration | (entry - duration date) / 365.25 years; duration date = symptom onset when available, else MS diagnosis date, else first DMT start |
| Baseline EDSS | Appendix B.2 (at-risk subset rules applied per outcome) |
| Parity | Count of live births before entry |
| Prior relapses (24 mo) | Count where (entry - 2 x 365.25 days) <= relapse date < entry |

A strict inequality on “before entry” matches the outcome convention of treating an on-entry relapse as follow-up, so it is not double-counted; parity uses the same strict inequality for consistency with the pregnancy exclusion and conception censor.

*G.2 Categorical Variables*

**Age category:** 1 = 40-44, 2 = 45-49, 3 = 50-54, 4 = 55-60.

**MS phenotype (SMSreg):** 1 = PPMS, 3 = RRMS [reference]. Records coded secondary progressive (raw code 2) are recoded to RRMS, so the baseline distinction is PPMS versus relapse-onset disease; Table 1 reports only PPMS and RRMS.

**Education (collapsed from original 1-7):** 1 = pre-secondary (1-2), 2 = secondary (3-5) [reference], 3 = university (6-7).

**Region of residence (NUTS-2):** 1 = Stockholm (SE11), 2 = Ostra Mellansverige (SE12), 3 = Smaland med oarna (SE21), 4 = Sydsverige (SE22), 5 = Vastsverige (SE23), 6 = Norra Mellansverige (SE31), 7 = Mellersta Norrland (SE32), 8 = Ovre Norrland (SE33). Derived from the first two digits of the kommun code; missing RTB region falls back to SMSreg treatment region.

**Calendar period:** 1 = 2006-2009, 2 = 2010-2014, 3 = 2015-2019, 4 = 2020-2024.

**Closest-year algorithm.** For time-varying registry data (education from LISA, region from RTB), the latest pre-entry yearly record is preferred; otherwise the earliest post-entry record is used.

### Appendix H. Missing Data Handling

Complete after exclusions: date of birth, sex, MS phenotype, MS diagnosis date (first-DMT proxy when missing), entry/exit dates, baseline EDSS. Count variables with no records are recoded to 0 (parity, prior relapses total and 24-month). First DMT class is recoded to 0 (No DMT) when missing.

Before imputation, education was missing in 45/7,623 (0.59%) and country of birth in <5/7,623. Education and country of birth are jointly imputed via chained equations (augmented multinomial logistic for education, augmented logistic for foreign birth). Predictors: age category, total births, calendar period, NUTS-2 region, HRT status, EDSS milestone indicators, and recurring-relapse indicator. Burn-in 20 iterations, one imputed draw retained, seed 1235813; the single draw is carried forward without multiple-imputation pooling.

**Limitation.** The single-draw approach does not propagate imputation uncertainty, so standard errors for education and country-of-birth coefficients may be slightly underestimated.

### Appendix I. Statistical Analysis

#### *I.1 Primary Analysis Models*

The primary framework is Cox proportional-hazards regression with time-varying exposure. Each exposure approach uses a separate analytic dataset; both share identical person-time (Appendix C.8).

| Specification | Exposure | Adjustment | Stratification |
|---|---|---|---|
| EDSS 4.0 / 6.0, binary | hrt_tv | TV DMT class, TV age (continuous), MS phenotype, disease duration, education, foreign birth, calendar year of entry (continuous), NUTS-2 region | Baseline EDSS |
| EDSS 4.0 / 6.0, dose | hrt_dosecat | Same | Baseline EDSS |

| Specification | Exposure | Adjustment | Stratification |
|---|---|---|---|
| Relapse, binary | hrt_tv | Same + baseline EDSS as covariate | Prior relapses (24 mo) |
| Relapse, dose | hrt_dosecat | Same + baseline EDSS as covariate | Prior relapses (24 mo) |

For EDSS outcomes the baseline hazard is stratified by baseline EDSS; for relapses it is stratified by prior-relapse count, with baseline EDSS as a covariate. These variables were moved into stratification because Schoenfeld testing (Appendix I.2) indicated non-proportional hazards when they entered as covariates; region was moved from stratification to covariate. Parity is not in the final specification. Cluster-robust standard errors on the person identifier account for within-person correlation from time-varying splitting. Reported sample sizes are distinct subjects, not person-time rows.

*I.2 Proportional Hazards Assessment*

The Schoenfeld residual test (robust variance) was applied to the binary HRT models.

| Outcome | Global chi-squared | df | Global p | HRT p |
|---|---|---|---|---|
| Sustained EDSS 4.0 | 26.47 | 18 | 0.09 | 0.65 |
| Sustained EDSS 6.0 | 13.14 | 18 | 0.78 | 0.22 |
| Recurring relapse | 202.97 | 19 | <0.001 | 0.31 |

The global test did not reject proportionality for EDSS outcomes. For relapses it was significant, driven by calendar year of entry, time-varying age, baseline EDSS, and region indicators rather than HRT. The HRT exposure term did not violate proportionality in any model, so the Cox specification is retained. The entry-age-50 sensitivity cohort showed the same pattern.

*I.3 Simon-Makuch Failure Curves*

Unadjusted Simon-Makuch failure curves (the Kaplan-Meier estimator extended to a time-varying exposure, in which each woman contributes to the current-use curve while using HRT and to the no-use curve otherwise) are plotted by time-varying binary HRT status for sustained EDSS 4.0 and 6.0 (panels A and B), for the primary age-40+ cohort (Figure 2) and the entry-age-50 sensitivity cohort (Figure S2). Panel C counts every recurring relapse, with women remaining at risk after each one, so it shows how relapses accumulate over time rather than the probability of a relapse.

*I.4 Entry-Age-50 Sensitivity Analysis*

Because the age-40-60 cohort mixes pre- and post-menopausal women, the unexposed group partially conflates HRT non-use with retained endogenous estrogen. This post-hoc analysis reruns the full pipeline with the entry-age floor raised from 40 to 50, holding all other inclusion, exclusion, exposure, and outcome rules constant. Study entry is recalculated as the latest of the 50th-birthday anniversary, MS diagnosis date, and 1 January 2006, again using the birthday-anniversary age calculation. The same Cox specifications (Appendix I.1) and at-risk EDSS rules apply. Binary and dose-category models are repeated for sustained EDSS 4.0, EDSS 6.0, and recurring relapses. The hysterectomy-excluded sensitivity analysis from earlier protocol versions is not included; prior hysterectomy is retained as a Table 1 descriptor.

*I.5 Output Tables and Figures*

| **Item** | **Content** | **Cohort** |
|---|---|---|
| Table 1 / S1 | Baseline characteristics by HRT use (binary) | Age 40+ primary / entry-age-50 |
| Table 2 / S2 | Events, person-years, unadjusted rates, adjusted HRs (both approaches) | Age 40+ primary / entry-age-50 |
| Table S3 / S4 | Full adjusted Cox model, binary / dose categories | Age 40+ primary |
| Table S5 / S6 | Full adjusted Cox model, binary / dose categories | Entry-age-50 |
| Table S7 / S8 | Events, rates, adjusted HRs (both approaches) | RRMS subgroup, age 40+ / entry-age-50 |
| Figure 1 / S1 | Sequential exclusion flowchart, starting from women alive and resident in Sweden at cohort entry | Age 40+ primary / entry-age-50 |
| Figure 2 / S2 | Unadjusted Simon-Makuch failure curves for sustained EDSS 4.0 and 6.0 (A, B) and cumulative recurring relapses (C) by HRT status | Age 40+ primary / entry-age-50 |

*I.6 Relapsing-Remitting MS Subgroup Analysis*

To assess whether results were driven by the primary progressive subgroup, the primary age-40+ analysis was repeated restricted to women with relapsing-remitting MS at diagnosis, and likewise for the entry-age-50 cohort. The phenotype term is dropped (collinear under restriction); all other covariates, stratification, and options are identical to the primary analysis. Results appear in Tables S7 (primary) and S8 (entry-age-50).

**Table S1.** Baseline characteristics by HRT use among the entry-age-50 sensitivity cohort

| No. (Column %) or Median (Q1, Q3) | Total N=4,725 | Never N=4,066 | HRT Use N=659 |
|---|---|---|---|
| Age | 50 (50, 52) | 50 (50, 53) | 50 (50, 50) |
| Age at baseline | | | |
| 50-54 | 3,942 (83) | 3,316 (82) | 626 (95) |
| 55-60 | 783 (17) | 750 (18) | 33 (5) |
| MS Phenotype at Diagnosis | | | |
| Primary Progressive | 354 (7) | 317 (8) | 37 (6) |
| Relapsing-Remitting | 4,371 (93) | 3,749 (92) | 622 (94) |
| Disease duration at baseline (years) | 14 (7, 22) | 14 (7, 22) | 14 (7, 21) |
| Highest education level | | | |
| Pre-secondary | 461 (10) | 415 (10) | 46 (7) |
| Secondary | 3,056 (65) | 2,654 (65) | 402 (61) |
| University Degree | 1,208 (26) | 997 (25) | 211 (32) |
| Born abroad | 434 (9) | 378 (9) | 56 (8) |
| Total number of births (parity) | | | |
| 0 | 1,028 (22) | 921 (23) | 107 (16) |
| 1 | 859 (18) | 728 (18) | 131 (20) |
| 2 | 1,904 (40) | 1,600 (39) | 304 (46) |
| 3 | 722 (15) | 633 (16) | 89 (14) |
| 4+ | 212 (4) | 184 (5) | 28 (4) |
| EDSS Baseline | | | |
| 0-3.5 | 3,253 (69) | 2,728 (67) | 525 (80) |
| 4-5.5 | 577 (12) | 521 (13) | 56 (8) |
| 6+ | 895 (19) | 817 (20) | 78 (12) |
| Number of relapses in 24 months before baseline | | | |
| 0 | 4,133 (87) | 3,554 (87) | 579 (88) |
| 1 | 455 (10) | 394 (10) | 61 (9) |
| 2 | 109 (2) | – | – |
| 3+ | 28 (1) | – | <5 |
| Residence region | | | |
| Stockholm | 1,143 (24) | 959 (24) | 184 (28) |
| Ostra Mellansverige | 879 (19) | 755 (19) | 124 (19) |
| Smaland med oarna | 307 (6) | 272 (7) | 35 (5) |
| Sydsverige | 588 (12) | 511 (13) | 77 (12) |
| Vastsverige | 935 (20) | 795 (20) | 140 (21) |
| Norra Mellansverige | 468 (10) | 411 (10) | 57 (9) |
| Mellersta Norrland | 172 (4) | 150 (4) | 22 (3) |
| Ovre Norrland | 233 (5) | 213 (5) | 20 (3) |
| Calendar Period | | | |
| 2006-2009 | 1,736 (37) | 1,589 (39) | 147 (22) |
| 2010-2014 | 1,040 (22) | 916 (23) | 124 (19) |
| 2015-2019 | 1,108 (23) | 890 (22) | 218 (33) |
| 2020-2024 | 841 (18) | 671 (17) | 170 (26) |
| Prior hysterectomy without oophorectomy | 140 (3) | 111 (3) | 29 (4) |
| First DMT during study period | | | |
| None | 1,219 (26) | 1,073 (26) | 146 (22) |
| Platform/Moderate | 2,120 (45) | 1,836 (45) | 284 (43) |
| High-efficacy | 350 (7) | 285 (7) | 65 (10) |
| Anti-CD20 | 1,036 (22) | 872 (21) | 164 (25) |
| Years of followup | 11 (7, 17) | 12 (7, 17) | 9 (6, 14) |

*Note: <5, count below the registry's disclosure threshold. En-dashed cells (–) are suppressed so that this count cannot be recovered by subtraction from the row or column totals.*

**Table S2.** HRT events, rates, and adjusted hazard ratios among the entry-age-50 sensitivity cohort

| | Sustained EDSS 4 | | | | | Sustained EDSS 6 | | | | | Recurring Relapse | | | | |
|---|---|---|---|---|---|---|---|---|---|---|---|---|---|---|---|
| **Exposure** | **Events** | **Person-Years (PY)** | **Per 1,000 PY (95% CI)** | **aHR (95% CI)** | **p-value** | **Events** | **Person-Years (PY)** | **Per 1,000 PY (95% CI)** | **aHR (95% CI)** | **p-value** | **Events** | **Person-Years (PY)** | **Per 1,000 PY (95% CI)** | **aHR (95% CI)** | **p-value** |
| Binary HRT | | | | | | | | | | | | | | | |
| None | 853 | 27,666 | 30.8 (28.8, 33.0) | Reference | | 772 | 35,772 | 21.6 (20.1, 23.2) | Reference | | 1,005 | 52,747 | 19.1 (17.9, 20.3) | Reference | |
| Current HRT use | 21 | 1,370 | 15.3 (10.0, 23.5) | 0.62 (0.40, 0.95) | 0.029 | 20 | 1,564 | 12.8 (8.2, 19.8) | 0.90 (0.57, 1.42) | 0.65 | 33 | 1,885 | 17.5 (12.4, 24.6) | 0.95 (0.60, 1.52) | 0.84 |
| Estrogen Dose Category | | | | | | | | | | | | | | | |
| None | 853 | 27,666 | 30.8 (28.8, 33.0) | Reference | | 772 | 35,772 | 21.6 (20.1, 23.2) | Reference | | 1,005 | 52,747 | 19.1 (17.9, 20.3) | Reference | |
| Low (<0.5mg) | – | – | – | 0.72 (0.18, 2.85) | 0.64 | – | – | – | 0.36 (0.04, 2.91) | 0.34 | – | – | – | 0.69 (0.18, 2.61) | 0.58 |
| Medium (0.5-1.5mg) | 14 | 1,060 | 13.2 (7.8, 22.3) | 0.55 (0.33, 0.94) | 0.029 | 14 | 1,198 | 11.7 (6.9, 19.7) | 0.93 (0.55, 1.57) | 0.80 | 21 | 1,396 | 15.0 (9.8, 23.1) | 0.86 (0.51, 1.44) | 0.56 |
| High (>1.5mg) | – | – | – | 0.81 (0.35, 1.89) | 0.63 | – | – | – | 1.12 (0.47, 2.70) | 0.79 | – | – | – | 1.38 (0.61, 3.13) | 0.43 |

*Note: HRT, Hormone Replacement Therapy; aHR, adjusted hazard ratio; CI, confidence interval. All models adjusted for age, calendar year, education, foreign country of birth, MS phenotype, disease duration, region of residence, and time-varying DMT class. Relapse model adjusts additionally for baseline EDSS. EDSS outcomes stratify baseline hazards by baseline EDSS. Relapse model stratifies baseline hazards by number of relapses in 24 months prior to baseline. Model standard errors clustered by patient id. En-dashed cells (–) are suppressed to comply with registry privacy restrictions because the underlying event count falls below the registry's disclosure threshold; within the estrogen-dose categories a further cell is suppressed where necessary to prevent a suppressed count from being recovered by subtraction from the published total.*

**Table S3.** Complete adjusted model results for binary HRT exposure (primary age 40+ cohort)

| | Sustained EDSS 4+ | | | Sustained EDSS 6+ | | | Recurring Relapses | | |
|---|---|---|---|---|---|---|---|---|---|
| | **HR** | **95% CI** | **p-value** | **HR** | **95% CI** | **p-value** | **HR** | **95% CI** | **p-value** |
| HRT (vs. None) | 0.70 | (0.51, 0.96) | 0.028 | 0.80 | (0.55, 1.16) | 0.24 | 1.00 | (0.74, 1.36) | 0.98 |
| Time-Varying DMT | | | | | | | | | |
| None | | *Reference* | | | *Reference* | | | *Reference* | |
| Platform/Moderate | 0.97 | (0.84, 1.12) | 0.66 | 0.77 | (0.65, 0.90) | 0.002 | 1.11 | (1.00, 1.23) | 0.056 |
| High-efficacy | 1.37 | (1.09, 1.73) | 0.006 | 1.13 | (0.88, 1.46) | 0.34 | 0.86 | (0.72, 1.03) | 0.10 |
| Anti-CD20 | 1.46 | (1.22, 1.74) | <0.001 | 1.42 | (1.17, 1.72) | <0.001 | 0.46 | (0.38, 0.56) | <0.001 |
| Age (time-varying) | 1.02 | (1.01, 1.03) | <0.001 | 1.03 | (1.02, 1.04) | <0.001 | 0.96 | (0.95, 0.96) | <0.001 |
| MS Phenotype at Diagnosis | | | | | | | | | |
| Primary Progressive | 1.94 | (1.56, 2.40) | <0.001 | 1.73 | (1.40, 2.13) | <0.001 | 0.39 | (0.28, 0.56) | <0.001 |
| Relapsing-Remitting | | *Reference* | | | *Reference* | | | *Reference* | |
| Disease duration at baseline (years) | 1.00 | (1.00, 1.01) | 0.29 | 1.00 | (1.00, 1.01) | 0.49 | 1.00 | (0.99, 1.00) | 0.15 |
| Highest education level | | | | | | | | | |
| Pre-secondary | 1.01 | (0.84, 1.21) | 0.92 | 1.00 | (0.83, 1.20) | >0.99 | 0.90 | (0.74, 1.10) | 0.31 |
| Secondary | | *Reference* | | | *Reference* | | | *Reference* | |
| University Degree | 0.75 | (0.66, 0.86) | <0.001 | 0.89 | (0.77, 1.03) | 0.11 | 1.01 | (0.90, 1.13) | 0.87 |
| Born abroad | 1.37 | (1.16, 1.62) | <0.001 | 1.31 | (1.07, 1.61) | 0.009 | 1.00 | (0.85, 1.18) | 0.97 |
| Year of study entry | 0.96 | (0.95, 0.98) | <0.001 | 0.96 | (0.94, 0.98) | <0.001 | 0.90 | (0.89, 0.92) | <0.001 |
| Residence region | | | | | | | | | |
| Stockholm | | *Reference* | | | *Reference* | | | *Reference* | |
| Ostra Mellansverige | 0.90 | (0.77, 1.07) | 0.23 | 1.16 | (0.96, 1.39) | 0.12 | 1.40 | (1.21, 1.61) | <0.001 |
| Smaland med oarna | 0.63 | (0.48, 0.82) | <0.001 | 0.78 | (0.58, 1.04) | 0.095 | 1.74 | (1.43, 2.11) | <0.001 |
| Sydsverige | 0.89 | (0.73, 1.07) | 0.21 | 1.17 | (0.96, 1.44) | 0.12 | 1.11 | (0.94, 1.31) | 0.23 |
| Vastsverige | 0.75 | (0.64, 0.88) | <0.001 | 0.89 | (0.75, 1.07) | 0.22 | 0.88 | (0.76, 1.02) | 0.091 |
| Norra Mellansverige | 0.88 | (0.72, 1.07) | 0.20 | 1.13 | (0.90, 1.42) | 0.30 | 1.29 | (1.08, 1.54) | 0.004 |
| Mellersta Norrland | 0.74 | (0.55, 1.00) | 0.049 | 0.87 | (0.61, 1.24) | 0.45 | 0.75 | (0.53, 1.08) | 0.12 |
| Ovre Norrland | 0.93 | (0.74, 1.17) | 0.52 | 1.01 | (0.77, 1.32) | 0.93 | 0.91 | (0.71, 1.16) | 0.44 |
| EDSS at baseline | | | | | | | 0.94 | (0.92, 0.97) | <0.001 |
| Subjects | | 5,908 | | | 6,611 | | | 7,623 | |

**Table S4.** Complete adjusted model results for estrogen dose categories (primary age 40+ cohort)

| | Sustained EDSS 4+ | | | Sustained EDSS 6+ | | | Recurring Relapses | | |
|---|---|---|---|---|---|---|---|---|---|
| | **HR** | **95% CI** | **p-value** | **HR** | **95% CI** | **p-value** | **HR** | **95% CI** | **p-value** |
| Estrogen Dose Category | | | | | | | | | |
| None | | *Reference* | | | *Reference* | | | *Reference* | |
| Low (<0.5mg) | 0.97 | (0.33, 2.86) | 0.96 | 1.38 | (0.47, 4.09) | 0.56 | 0.73 | (0.26, 2.07) | 0.55 |
| Medium (0.5-1.5mg) | 0.69 | (0.47, 0.99) | 0.046 | 0.80 | (0.52, 1.23) | 0.31 | 0.96 | (0.66, 1.39) | 0.83 |
| High (>1.5mg) | 0.68 | (0.34, 1.35) | 0.27 | 0.63 | (0.28, 1.43) | 0.27 | 1.17 | (0.72, 1.92) | 0.52 |
| Time-Varying DMT | | | | | | | | | |
| None | | *Reference* | | | *Reference* | | | *Reference* | |
| Platform/Moderate | 0.97 | (0.84, 1.12) | 0.67 | 0.76 | (0.64, 0.90) | 0.002 | 1.11 | (1.00, 1.23) | 0.056 |
| High-efficacy | 1.37 | (1.09, 1.73) | 0.006 | 1.13 | (0.88, 1.46) | 0.34 | 0.86 | (0.72, 1.03) | 0.11 |
| Anti-CD20 | 1.46 | (1.22, 1.74) | <0.001 | 1.41 | (1.17, 1.72) | <0.001 | 0.46 | (0.38, 0.56) | <0.001 |
| Age (time-varying) | 1.02 | (1.01, 1.03) | <0.001 | 1.03 | (1.02, 1.04) | <0.001 | 0.96 | (0.95, 0.96) | <0.001 |
| MS Phenotype at Diagnosis | | | | | | | | | |
| Primary Progressive | 1.94 | (1.56, 2.40) | <0.001 | 1.72 | (1.40, 2.12) | <0.001 | 0.39 | (0.28, 0.56) | <0.001 |
| Relapsing-Remitting | | *Reference* | | | *Reference* | | | *Reference* | |
| Disease duration at baseline (years) | 1.00 | (1.00, 1.01) | 0.29 | 1.00 | (1.00, 1.01) | 0.51 | 1.00 | (0.99, 1.00) | 0.15 |
| Highest education level | | | | | | | | | |
| Pre-secondary | 1.01 | (0.84, 1.21) | 0.92 | 1.00 | (0.83, 1.21) | 0.99 | 0.90 | (0.74, 1.10) | 0.31 |
| Secondary | | *Reference* | | | *Reference* | | | *Reference* | |
| University Degree | 0.75 | (0.66, 0.86) | <0.001 | 0.89 | (0.77, 1.03) | 0.12 | 1.01 | (0.90, 1.13) | 0.86 |
| Born abroad | 1.37 | (1.16, 1.62) | <0.001 | 1.31 | (1.07, 1.61) | 0.008 | 1.00 | (0.85, 1.18) | 0.97 |
| Year of study entry | 0.96 | (0.95, 0.98) | <0.001 | 0.96 | (0.94, 0.98) | <0.001 | 0.90 | (0.89, 0.92) | <0.001 |
| Residence region | | | | | | | | | |
| Stockholm | | *Reference* | | | *Reference* | | | *Reference* | |
| Ostra Mellansverige | 0.90 | (0.77, 1.07) | 0.23 | 1.16 | (0.96, 1.39) | 0.12 | 1.39 | (1.21, 1.61) | <0.001 |
| Smaland med oarna | 0.63 | (0.48, 0.82) | <0.001 | 0.78 | (0.58, 1.05) | 0.095 | 1.74 | (1.43, 2.11) | <0.001 |
| Sydsverige | 0.89 | (0.73, 1.07) | 0.21 | 1.18 | (0.96, 1.44) | 0.12 | 1.11 | (0.94, 1.31) | 0.23 |
| Vastsverige | 0.75 | (0.64, 0.88) | <0.001 | 0.89 | (0.75, 1.07) | 0.22 | 0.88 | (0.76, 1.02) | 0.089 |
| Norra Mellansverige | 0.88 | (0.72, 1.07) | 0.20 | 1.13 | (0.90, 1.42) | 0.30 | 1.29 | (1.08, 1.54) | 0.004 |
| Mellersta Norrland | 0.74 | (0.55, 1.00) | 0.049 | 0.87 | (0.61, 1.24) | 0.45 | 0.75 | (0.52, 1.07) | 0.12 |
| Ovre Norrland | 0.93 | (0.74, 1.17) | 0.52 | 1.01 | (0.77, 1.32) | 0.93 | 0.91 | (0.71, 1.16) | 0.44 |
| EDSS at baseline | | | | | | | 0.94 | (0.92, 0.97) | <0.001 |
| Subjects | | 5,908 | | | 6,611 | | | 7,623 | |

**Table S5.** Complete adjusted model results for binary HRT exposure among the entry-age-50 sensitivity cohort

| | Sustained EDSS 4+ | | | Sustained EDSS 6+ | | | Recurring Relapses | | |
|---|---|---|---|---|---|---|---|---|---|
| | **HR** | **95% CI** | **p-value** | **HR** | **95% CI** | **p-value** | **HR** | **95% CI** | **p-value** |
| HRT (vs. None) | 0.62 | (0.40, 0.95) | 0.029 | 0.90 | (0.57, 1.42) | 0.65 | 0.95 | (0.60, 1.52) | 0.84 |
| Time-Varying DMT | | | | | | | | | |
| None | | *Reference* | | | *Reference* | | | *Reference* | |
| Platform/Moderate | 1.12 | (0.93, 1.34) | 0.22 | 0.82 | (0.67, 1.00) | 0.050 | 1.10 | (0.93, 1.31) | 0.27 |
| High-efficacy | 1.29 | (0.92, 1.82) | 0.14 | 0.99 | (0.68, 1.45) | 0.96 | 0.89 | (0.63, 1.27) | 0.52 |
| Anti-CD20 | 1.57 | (1.27, 1.94) | <0.001 | 1.49 | (1.19, 1.86) | <0.001 | 0.49 | (0.36, 0.68) | <0.001 |
| Age (time-varying) | 1.03 | (1.01, 1.06) | 0.011 | 1.01 | (0.99, 1.04) | 0.33 | 0.94 | (0.91, 0.97) | <0.001 |
| MS Phenotype at Diagnosis | | | | | | | | | |
| Primary Progressive | 1.79 | (1.38, 2.31) | <0.001 | 1.57 | (1.24, 1.97) | <0.001 | 0.32 | (0.21, 0.49) | <0.001 |
| Relapsing-Remitting | | *Reference* | | | *Reference* | | | *Reference* | |
| Disease duration at baseline (years) | 1.00 | (0.99, 1.01) | 0.97 | 1.00 | (0.99, 1.01) | 0.58 | 0.98 | (0.98, 0.99) | <0.001 |
| Highest education level | | | | | | | | | |
| Pre-secondary | 1.03 | (0.82, 1.29) | 0.79 | 1.01 | (0.82, 1.26) | 0.91 | 0.94 | (0.70, 1.25) | 0.66 |
| Secondary | | *Reference* | | | *Reference* | | | *Reference* | |
| University Degree | 0.87 | (0.74, 1.02) | 0.092 | 0.86 | (0.73, 1.03) | 0.10 | 1.14 | (0.94, 1.37) | 0.18 |
| Born abroad | 1.40 | (1.12, 1.76) | 0.004 | 1.34 | (1.04, 1.74) | 0.025 | 1.05 | (0.80, 1.39) | 0.71 |
| Year of study entry | 0.98 | (0.96, 1.00) | 0.025 | 0.98 | (0.96, 1.00) | 0.016 | 0.89 | (0.87, 0.91) | <0.001 |
| Residence region | | | | | | | | | |
| Stockholm | | *Reference* | | | *Reference* | | | *Reference* | |
| Ostra Mellansverige | 0.77 | (0.63, 0.95) | 0.016 | 1.10 | (0.88, 1.37) | 0.40 | 1.49 | (1.16, 1.91) | 0.002 |
| Smaland med oarna | 0.55 | (0.40, 0.77) | <0.001 | 0.76 | (0.54, 1.08) | 0.12 | 1.78 | (1.31, 2.41) | <0.001 |
| Sydsverige | 0.81 | (0.63, 1.04) | 0.093 | 1.18 | (0.93, 1.49) | 0.18 | 1.26 | (0.94, 1.68) | 0.12 |
| Vastsverige | 0.66 | (0.54, 0.81) | <0.001 | 0.84 | (0.68, 1.05) | 0.13 | 0.92 | (0.71, 1.19) | 0.53 |
| Norra Mellansverige | 0.83 | (0.65, 1.06) | 0.14 | 1.19 | (0.91, 1.56) | 0.21 | 1.64 | (1.21, 2.21) | 0.001 |
| Mellersta Norrland | 0.63 | (0.44, 0.91) | 0.013 | 0.87 | (0.57, 1.34) | 0.53 | 0.65 | (0.37, 1.15) | 0.14 |
| Ovre Norrland | 1.00 | (0.75, 1.34) | >0.99 | 1.16 | (0.85, 1.58) | 0.35 | 0.99 | (0.67, 1.48) | 0.97 |
| EDSS at baseline | | | | | | | 0.95 | (0.91, 1.00) | 0.039 |
| Subjects | | 3,253 | | | 3,830 | | | 4,725 | |

**Table S6.** Complete adjusted model results for estrogen dose categories among the entry-age-50 sensitivity cohort

| | Sustained EDSS 4+ | | | Sustained EDSS 6+ | | | Recurring Relapses | | |
|---|---|---|---|---|---|---|---|---|---|
| | **HR** | **95% CI** | **p-value** | **HR** | **95% CI** | **p-value** | **HR** | **95% CI** | **p-value** |
| Estrogen Dose Category | | | | | | | | | |
| None | | *Reference* | | | *Reference* | | | *Reference* | |
| Low (<0.5mg) | 0.72 | (0.18, 2.85) | 0.64 | 0.36 | (0.04, 2.91) | 0.34 | 0.69 | (0.18, 2.61) | 0.58 |
| Medium (0.5-1.5mg) | 0.55 | (0.33, 0.94) | 0.029 | 0.93 | (0.55, 1.57) | 0.80 | 0.86 | (0.51, 1.44) | 0.56 |
| High (>1.5mg) | 0.81 | (0.35, 1.89) | 0.63 | 1.12 | (0.47, 2.70) | 0.79 | 1.38 | (0.61, 3.13) | 0.43 |
| Time-Varying DMT | | | | | | | | | |
| None | | *Reference* | | | *Reference* | | | *Reference* | |
| Platform/Moderate | 1.12 | (0.93, 1.34) | 0.22 | 0.82 | (0.68, 1.00) | 0.055 | 1.10 | (0.93, 1.31) | 0.27 |
| High-efficacy | 1.29 | (0.92, 1.82) | 0.14 | 0.99 | (0.68, 1.45) | 0.98 | 0.89 | (0.63, 1.27) | 0.53 |
| Anti-CD20 | 1.57 | (1.27, 1.94) | <0.001 | 1.49 | (1.20, 1.86) | <0.001 | 0.49 | (0.36, 0.68) | <0.001 |
| Age (time-varying) | 1.03 | (1.01, 1.06) | 0.011 | 1.01 | (0.99, 1.04) | 0.34 | 0.94 | (0.91, 0.97) | <0.001 |
| MS Phenotype at Diagnosis | | | | | | | | | |
| Primary Progressive | 1.79 | (1.38, 2.31) | <0.001 | 1.58 | (1.25, 1.99) | <0.001 | 0.32 | (0.21, 0.49) | <0.001 |
| Relapsing-Remitting | | *Reference* | | | *Reference* | | | *Reference* | |
| Disease duration at baseline (years) | 1.00 | (0.99, 1.01) | 0.97 | 1.00 | (0.99, 1.01) | 0.59 | 0.98 | (0.98, 0.99) | <0.001 |
| Highest education level | | | | | | | | | |
| Pre-secondary | 1.03 | (0.82, 1.29) | 0.78 | 1.01 | (0.81, 1.25) | 0.93 | 0.94 | (0.70, 1.26) | 0.66 |
| Secondary | | *Reference* | | | *Reference* | | | *Reference* | |
| University Degree | 0.87 | (0.75, 1.02) | 0.095 | 0.86 | (0.72, 1.03) | 0.095 | 1.14 | (0.95, 1.37) | 0.16 |
| Born abroad | 1.40 | (1.11, 1.76) | 0.004 | 1.34 | (1.04, 1.74) | 0.025 | 1.06 | (0.80, 1.39) | 0.70 |
| Year of study entry | 0.98 | (0.96, 1.00) | 0.026 | 0.97 | (0.96, 1.00) | 0.015 | 0.89 | (0.87, 0.91) | <0.001 |
| Residence region | | | | | | | | | |
| Stockholm | | *Reference* | | | *Reference* | | | *Reference* | |
| Ostra Mellansverige | 0.77 | (0.63, 0.95) | 0.015 | 1.09 | (0.88, 1.36) | 0.42 | 1.48 | (1.15, 1.90) | 0.002 |
| Smaland med oarna | 0.55 | (0.40, 0.77) | <0.001 | 0.76 | (0.54, 1.07) | 0.12 | 1.78 | (1.31, 2.41) | <0.001 |
| Sydsverige | 0.80 | (0.63, 1.04) | 0.091 | 1.17 | (0.93, 1.49) | 0.18 | 1.25 | (0.94, 1.67) | 0.12 |
| Vastsverige | 0.66 | (0.54, 0.81) | <0.001 | 0.84 | (0.68, 1.05) | 0.13 | 0.92 | (0.71, 1.19) | 0.52 |
| Norra Mellansverige | 0.83 | (0.65, 1.06) | 0.14 | 1.19 | (0.90, 1.56) | 0.22 | 1.63 | (1.21, 2.21) | 0.001 |
| Mellersta Norrland | 0.63 | (0.44, 0.91) | 0.013 | 0.87 | (0.57, 1.33) | 0.53 | 0.65 | (0.37, 1.15) | 0.14 |
| Ovre Norrland | 1.00 | (0.75, 1.34) | >0.99 | 1.16 | (0.85, 1.58) | 0.36 | 0.99 | (0.67, 1.47) | 0.96 |
| EDSS at baseline | | | | | | | 0.95 | (0.91, 1.00) | 0.039 |
| Subjects | | 3,253 | | | 3,830 | | | 4,725 | |

**Table S7.** HRT events, rates, and adjusted hazard ratios among relapsing-remitting MS subgroup (primary age 40+ cohort)

| | Sustained EDSS 4 | | | | | Sustained EDSS 6 | | | | | Recurring Relapse | | | | |
|---|---|---|---|---|---|---|---|---|---|---|---|---|---|---|---|
| **Exposure** | **Events** | **Person-Years (PY)** | **Per 1,000 PY (95% CI)** | **aHR (95% CI)** | **p-value** | **Events** | **Person-Years (PY)** | **Per 1,000 PY (95% CI)** | **aHR (95% CI)** | **p-value** | **Events** | **Person-Years (PY)** | **Per 1,000 PY (95% CI)** | **aHR (95% CI)** | **p-value** |
| Binary HRT | | | | | | | | | | | | | | | |
| None | 1,256 | 55,869 | 22.5 (21.3, 23.8) | Reference | | 965 | 67,769 | 14.2 (13.4, 15.2) | Reference | | 2,917 | 87,749 | 33.2 (32.1, 34.5) | Reference | |
| Current HRT use | – | – | – | 0.71 (0.51, 0.97) | 0.033 | – | – | – | 0.78 (0.53, 1.16) | 0.22 | 67 | 3,373 | 19.9 (15.6, 25.2) | 1.02 (0.75, 1.38) | 0.91 |
| Estrogen Dose Category | | | | | | | | | | | | | | | |
| None | 1,256 | 55,869 | 22.5 (21.3, 23.8) | Reference | | 965 | 67,769 | 14.2 (13.4, 15.2) | Reference | | 2,917 | 87,749 | 33.2 (32.1, 34.5) | Reference | |
| Low (<0.5mg) | – | – | – | 0.97 (0.33, 2.87) | 0.96 | – | – | – | 1.92 (0.75, 4.93) | 0.18 | – | – | – | 0.75 (0.26, 2.11) | 0.58 |
| Medium (0.5-1.5mg) | – | – | – | 0.68 (0.47, 1.00) | 0.050 | – | – | – | 0.76 (0.47, 1.21) | 0.24 | 43 | 2,401 | 17.9 (13.3, 24.2) | 0.97 (0.67, 1.41) | 0.88 |
| High (>1.5mg) | – | – | – | 0.70 (0.35, 1.39) | 0.31 | – | – | – | 0.57 (0.23, 1.43) | 0.23 | – | – | – | 1.20 (0.73, 1.96) | 0.48 |

*Note: HRT, Hormone Replacement Therapy; aHR, adjusted hazard ratio; CI, confidence interval. All models adjusted for age, calendar year, education, foreign country of birth, disease duration, region of residence, and time-varying DMT class. Relapse model adjusts additionally for baseline EDSS. EDSS outcomes stratify baseline hazards by baseline EDSS. Relapse model stratifies baseline hazards by number of relapses in 24 months prior to baseline. Model standard errors clustered by patient id. En-dashed cells (–) are suppressed to comply with registry privacy restrictions because the underlying event count falls below the registry's disclosure threshold; within the estrogen-dose categories a further cell is suppressed where necessary to prevent a suppressed count from being recovered by subtraction from the published total. In this table, events, person-years and rates for current HRT use and the medium dose are also suppressed for the EDSS outcomes, because subtracting them from Table 2 would reveal fewer than five events among women with primary progressive MS.*

**Table S8.** HRT events, rates, and adjusted hazard ratios among relapsing-remitting MS subgroup (entry-age-50 sensitivity cohort)

| | **Sustained EDSS 4** | | | | | **Sustained EDSS 6** | | | | | **Recurring Relapse** | | | | |
|---|---|---|---|---|---|---|---|---|---|---|---|---|---|---|---|
| **Exposure** | **Events** | **Person-Years (PY)** | **Per 1,000 PY (95% CI)** | **aHR (95% CI)** | **p-value** | **Events** | **Person-Years (PY)** | **Per 1,000 PY (95% CI)** | **aHR (95% CI)** | **p-value** | **Events** | **Person-Years (PY)** | **Per 1,000 PY (95% CI)** | **aHR (95% CI)** | **p-value** |
| Binary HRT | | | | | | | | | | | | | | | |
| None | 775 | 26,584 | 29.2 (27.2, 31.3) | Reference | | 668 | 33,923 | 19.7 (18.3, 21.2) | Reference | | 979 | 48,256 | 20.3 (19.1, 21.6) | Reference | |
| Current HRT use | 21 | 1,357 | 15.5 (10.1, 23.7) | 0.64 (0.41, 0.98) | 0.041 | 20 | 1,527 | 13.1 (8.4, 20.3) | 1.07 (0.69, 1.67) | 0.77 | 33 | 1,823 | 18.1 (12.9, 25.5) | 0.97 (0.61, 1.54) | 0.88 |
| Estrogen Dose Category | | | | | | | | | | | | | | | |
| None | 775 | 26,584 | 29.2 (27.2, 31.3) | Reference | | 668 | 33,923 | 19.7 (18.3, 21.2) | Reference | | 979 | 48,256 | 20.3 (19.1, 21.6) | Reference | |
| Low (<0.5mg) | – | – | – | 0.72 (0.18, 2.89) | 0.65 | – | – | – | 0.62 (0.09, 4.27) | 0.63 | – | – | – | 0.71 (0.19, 2.73) | 0.62 |
| Medium (0.5-1.5mg) | 14 | 1,047 | 13.4 (7.9, 22.6) | 0.58 (0.34, 0.98) | 0.042 | 14 | 1,169 | 12.0 (7.1, 20.2) | 1.07 (0.64, 1.78) | 0.80 | 21 | 1,359 | 15.4 (10.1, 23.7) | 0.87 (0.52, 1.46) | 0.59 |
| High (>1.5mg) | – | – | – | 0.83 (0.36, 1.93) | 0.66 | – | – | – | 1.26 (0.52, 3.07) | 0.61 | – | – | – | 1.39 (0.61, 3.14) | 0.43 |

*Note: HRT, Hormone Replacement Therapy; aHR, adjusted hazard ratio; CI, confidence interval. All models adjusted for age, calendar year, education, foreign country of birth, disease duration, region of residence, and time-varying DMT class. Relapse model adjusts additionally for baseline EDSS. EDSS outcomes stratify baseline hazards by baseline EDSS. Relapse model stratifies baseline hazards by number of relapses in 24 months prior to baseline. Model standard errors clustered by patient id. En-dashed cells (–) are suppressed to comply with registry privacy restrictions because the underlying event count falls below the registry's disclosure threshold; within the estrogen-dose categories a further cell is suppressed where necessary to prevent a suppressed count from being recovered by subtraction from the published total.*

**Figure S1.** Sequential exclusion flowchart, starting from women alive and resident in Sweden at cohort entry (entry-age-50 sensitivity cohort).

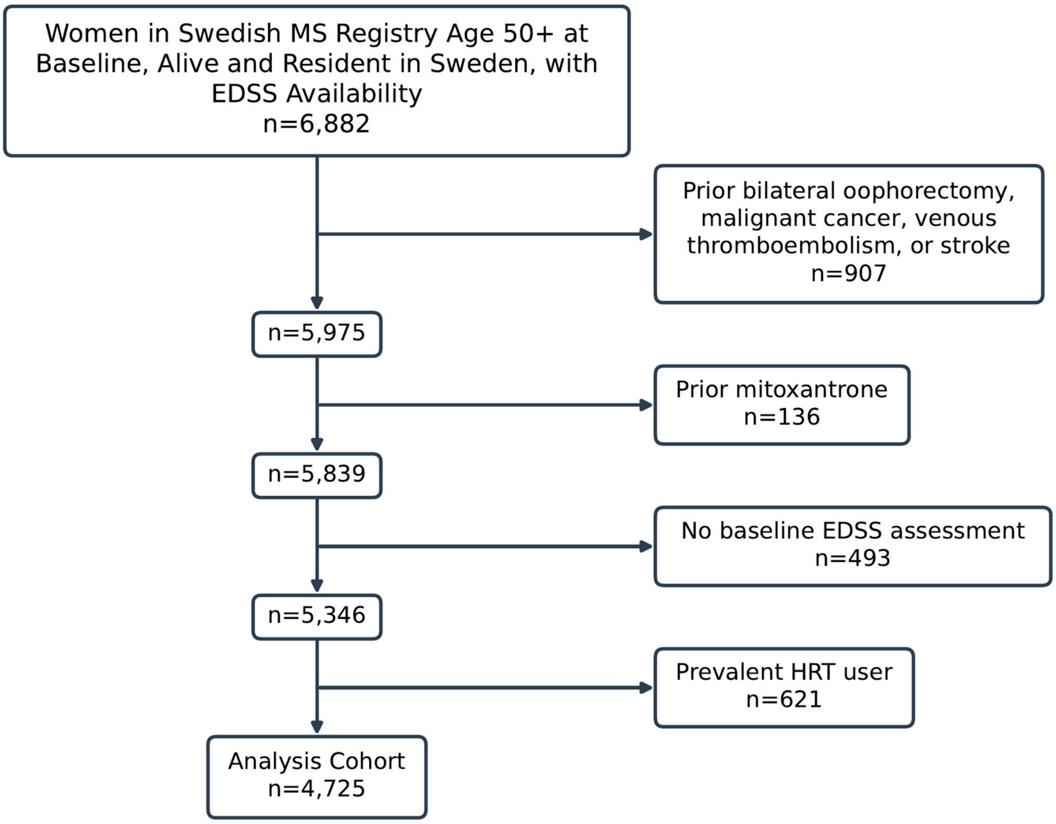

**Figure S2.** Unadjusted Simon-Makuch failure curves for sustained EDSS 4.0 and 6.0 (A, B) and cumulative recurring relapses (C) by HRT status (entry-age-50 sensitivity cohort).

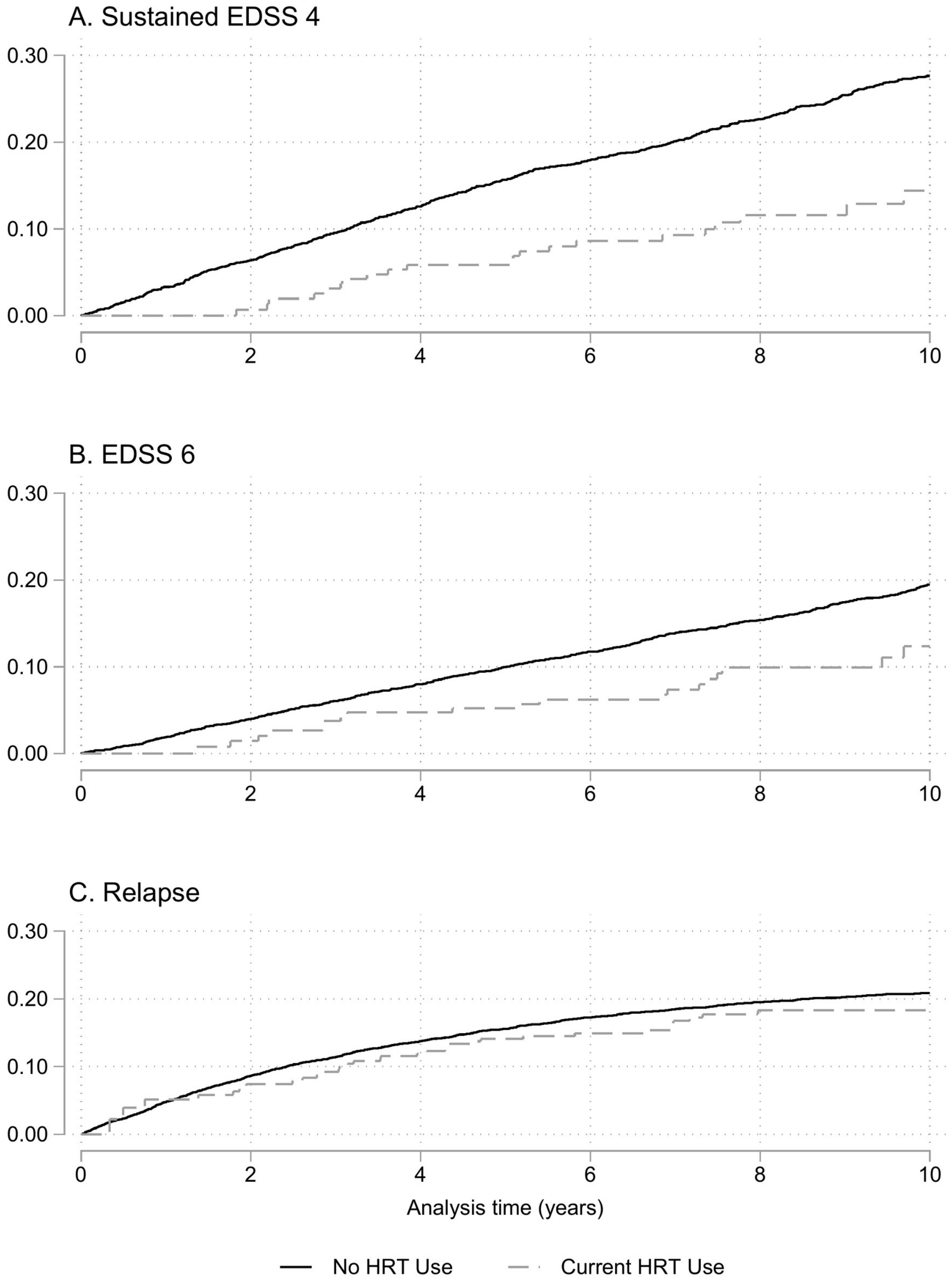

### RECORD checklist

**Manuscript:** Hormone Replacement Therapy and Disability Progression in Women with Multiple Sclerosis

Benchimol EI, et al. PLoS Med 2015;12(10):e1001885 (STROBE cohort items with RECORD extensions). All items are reported; exceptions are noted. Methods subsections: Study Design, Data Sources, Study Population, Exposure Definition, Outcomes, Covariates, Statistical Analysis, Sensitivity Analyses, Standard Protocol Approvals, Data Availability. App, Technical Appendix (Appendices A to I); S, Supplementary Table.

| Item | Topic | Where reported |
|---|---|---|
| 1 | Design in title; informative abstract | Abstract, Methods (design not stated in the title) |
| 1.1 | Data type and databases named | Abstract, Methods (registry-based; registers not named in the abstract); Methods, Data Sources |
| 1.2 | Region and time frame | Abstract, Methods (not in the title) |
| 1.3 | Linkage stated | Methods, Data Sources; App A.2 (not in the abstract) |
| 2 | Background and rationale | Introduction |
| 3 | Objectives | Abstract, Background; Introduction, last paragraph |
| 4 | Study design | Methods, Study Design |
| 5 | Setting and dates | Methods, Study Population; App B.2 |
| 6 | Eligibility and follow-up | Methods, Study Population; App B.1 to B.4, D.3 (no matching) |
| 6.1 | Codes and algorithms for the population | Methods, Study Population; App B.1, B.3, B.4 |
| 6.2 | Validation of codes | App A.2 (McDonald criteria field; no validation study of the selection algorithm cited) |
| 6.3 | Linkage flow diagram | Figure 1; Figure S1 (selection flowcharts; no linkage-level diagram) |
| 7 | Variable definitions | Methods, Exposure Definition, Outcomes, Covariates; App C, D, E, G |
| 7.1 | Codes for all variables | App B.4, C.1, E.1 |
| 8 | Data sources and measurement | Methods, Data Sources, Exposure Definition, Outcomes; App A.1, C.2 to C.7, D.1 |
| 9 | Bias | Methods, Study Design, Statistical Analysis, Sensitivity Analyses; App B.4; Discussion |

| Item | Topic | Where reported |
|---|---|---|
| 10 | Study size | Results, Study Population; Figure 1 (no sample size calculation) |
| 11 | Quantitative variables | Methods, Exposure Definition, Covariates; App C.8, G.1, G.2 |
| 12 | Statistical methods, interactions, missing data, follow-up, sensitivity analyses | Methods, Covariates, Statistical Analysis, Sensitivity Analyses; App H, I.1, I.2, I.4, I.6 (no interaction analyses) |
| 12.1 | Data access | Methods, Data Availability; App A.2 |
| 12.2 | Data cleaning | App B.2, C.2 to C.4, E.2 to E.4, F |
| 12.3 | Linkage method and quality | Methods, Data Sources; App A.2 (linkage quality not reported) |
| 13, 13.1 | Participants at each stage; selection | Results, Study Population; Figure 1; Figure S1 |
| 14 | Descriptive data, missing data, follow-up time | Table 1; S1; Results, Study Population; Methods, Covariates; App H |
| 15 | Outcome data | Results, Unadjusted Rates; Table 2; S2, S7, S8 |
| 16 | Main results, category boundaries, absolute measures | Results, Unadjusted Rates, Primary Analyses; Table 2; S3, S4; Methods, Exposure Definition |
| 17 | Other analyses | Results, Proportional Hazards Assessment, Sensitivity Analyses; S1, S2, S5 to S8; App I.2, I.4, I.6 |
| 18 | Key results | Discussion, first paragraph |
| 19, 19.1 | Limitations; implications of routinely collected data | Discussion, limitations paragraph |
| 20 | Interpretation | Discussion; conclusion paragraph |
| 21 | Generalisability | Discussion, conclusion paragraph |
| 22 | Funding | Funding statement |
| 22.1 | Protocol, data and code access | Methods, Data Availability (code on request; no protocol registered) |
